# POTENTIATION OF CORTICO-SPINAL OUTPUT VIA TARGETED ELECTRICAL STIMULATION OF THE MOTOR THALAMUS

**DOI:** 10.1101/2023.03.08.23286720

**Authors:** Jonathan C. Ho, Erinn M. Grigsby, Arianna Damiani, Lucy Liang, Josep-Maria Balaguer, Sridula Kallakuri, Jessica Barrios-Martinez, Vahagn Karapetyan, Daryl Fields, Peter C. Gerszten, T. Kevin Hitchens, Theodora Constantine, Gregory M. Adams, Donald J. Crammond, Marco Capogrosso, Jorge A. Gonzalez-Martinez, Elvira Pirondini

## Abstract

Cerebral white matter lesions prevent cortico-spinal descending inputs from effectively activating spinal motoneurons, leading to loss of motor control. However, in most cases, the damage to cortico-spinal axons is incomplete offering a potential target for new therapies aimed at improving volitional muscle activation. Here we hypothesized that, by engaging direct excitatory connections to cortico-spinal motoneurons, stimulation of the motor thalamus could facilitate activation of surviving cortico-spinal fibers thereby potentiating motor output. To test this hypothesis, we identified optimal thalamic targets and stimulation parameters that enhanced upper-limb motor evoked potentials and grip forces in anesthetized monkeys. This potentiation persisted after white matter lesions. We replicated these results in humans during intra-operative testing. We then designed a stimulation protocol that immediately improved voluntary grip force control in a patient with a chronic white matter lesion. Our results show that electrical stimulation targeting surviving neural pathways can improve motor control after white matter lesions.

## Main

Lesions of the cortico-spinal tract (CST), as a consequence of stroke, traumatic brain injury (TBI), brain tumors, or neurodegenerative disorders, disrupt communication between the cortex and lower motor centers leading to deficits and potential loss of function in face, upper or lower limb muscles^1^. The consequent upper-limb motor deficits significantly affect the quality of life of patients. It is estimated that approximately 10 million people in the United States^2,3^ alone live with such impairments.

In most cases though, damage to the CST is incomplete. Yet the spared excitatory descending connections are insufficient to fully activate the spinal motoneurons, leading to functional motor deficits^1^. Facilitation of the activation of the residual cortico-spinal axons could reestablish the missing excitation, restoring voluntary muscle control. We conjectured that this facilitation could be achieved by increasing the excitability of cortico-spinal neurons in the motor cortex, thereby increasing CST output, and consequently enhancing movements of the paretic limb^4,5^.

Stimulation of subcortical regions could be a viable approach to modulate the excitability of cortico-spinal neurons. Indeed, while certain mechanisms of action of deep brain stimulation (DBS) are still unclear, it is known that stimulation-induced action potentials within the subcortical targets propagate through axon terminals to recipient cortical areas modulating their neural activity^6^. Therefore, DBS can be used to increase excitability of restricted cortical areas by targeting subcortical regions that preferentially project excitatory axons to those areas.

For example, previous studies have demonstrated that stimulation of thalamic and subthalamic nuclei affect motor cortex excitability. Specifically, motor evoked potentials (MEPs) elicited by transcranial magnetic stimulation (TMS) were modulated when conditioned by stimulation of subcortical brain areas with direct (ventral thalamus) and indirect (globus pallidus pars interna, GPi and subthalamic nucleus, STN) projections to the motor cortex^7–11^.

Based on these results and the known anatomical evidence of direct connections to the motor cortex from the thalamic motor nuclei^8,12–15^, we hypothesized that stimulation of the motor thalamus could be tailored to facilitate motor cortex and CST activation and consequently improve motor deficits after lesions of the CST. However, identification of which nuclei of the motor thalamus should be targeted and which stimulation parameters would maximize both specificity and efficacy of thalamic stimulation is necessary to tailor DBS for treating upper-limb motor deficits.

Here, we overcame these scientific and technological challenges with a translational experimental framework that leveraged experiments in monkeys, i.e., the most relevant animal model for CST anatomy and function^1,16,17^, to verify mechanisms of action and identify optimal targets, and subsequently tested these findings in human patients. Specifically, we 1) identified the nucleus of the motor thalamus with dense, direct, and preferential excitatory connections to cortico-spinal neurons in primary motor cortex; 2) designed a series of electrophysiology experiments in monkeys and human patients to test whether stimulation of this nucleus could augment excitability of the motor cortex with high specificity to cause increased motor output; and 3) optimized stimulation parameters to maximize these effects. We then built on these findings to demonstrate that targeted stimulation of the motor thalamus increased muscle activation, strength, and force control after lesions of the CST in intraoperative monkeys and human experiments. Finally, we verified that these effects lead to improved functional performance during behavioral tasks in a subject with a chronic lesion of the CST and a DBS system implanted in the motor thalamus.

Overall, we demonstrated the mechanisms of action of motor thalamus stimulation and we optimized it for upper-limb paresis, paving the way towards a novel therapy for motor deficits after CST lesions.

## Results

### Identification of optimal target of stimulation within the motor thalamus

Here we aimed at increasing the excitability of cortico-spinal neurons within the primary motor cortex to potentiate motor output (**Fig 1a**). We therefore sought to identify the optimal stimulation target by localizing a subcortical region that has a high number of direct excitatory projections to the motor cortex. Based on previous anatomical evidence, we posited that the motor thalamus could be this optimal target^8,12,14,18^. In humans, the motor thalamus includes four nuclei that have different preferential projection targets: the ventral caudal (VC), the ventral intermediate (VIM), the ventral oral posterior (VOP), and the ventral anterior (VA) nucleus^19,20^. Thalamocortical fibers originating in the VC project preferentially to the somatosensory cortex, whereas those originating in the VIM/VOP connect to the motor cortex, and those in the VA to the pre-motor cortex and supplementary motor area (SMA). We first acquired and analyzed high-resolution diffusion magnetic resonance imaging (MRI) data using high-definition fiber tracking (HDFT) in monkeys (n=3) to confirm a similar anatomical organization, as previously shown with histological analysis^13–15^, and to identify the optimal target of stimulation. We focused our analysis on the three nuclei of the monkey motor thalamus: the ventral anterolateral (VAL), the ventral laterolateral (VLL), and the ventral posterolateral (VPL) nuclei^13–15,21–23^. We reconstructed all the likely axonal pathways between these nuclei and the somatosensory, motor, and premotor cortical areas and quantified the relative strength of these connections by calculating the volume of thalamocortical projections from each nucleus to each cortical region normalized by the total volume of fibers (**Fig. 1b**). We confirmed a clear functional and somatotopic organization of the primate thalamic nuclei that parallel the organization of the human thalamocortical connections, with axonal projections going preferentially towards the somatosensory cortex from the posterior motor thalamus (VPL) and preferentially towards motor and pre-motor cortices from the anterior motor thalamus (i.e., VLL and VAL, respectively). Overall these results helped to define the optimal stimulation target. Indeed, using selectivity detection analysis among the three nuclei, we found that the VLL nucleus, which corresponds to the human VIM/VOP nucleus, had the greatest selectivity of the projections towards M1 (VPL: 0.67, VLL: 0.89, VAL: 0.33). We, therefore, considered the VLL nucleus the optimal target to potentiate motor output.

**Fig. 1:**
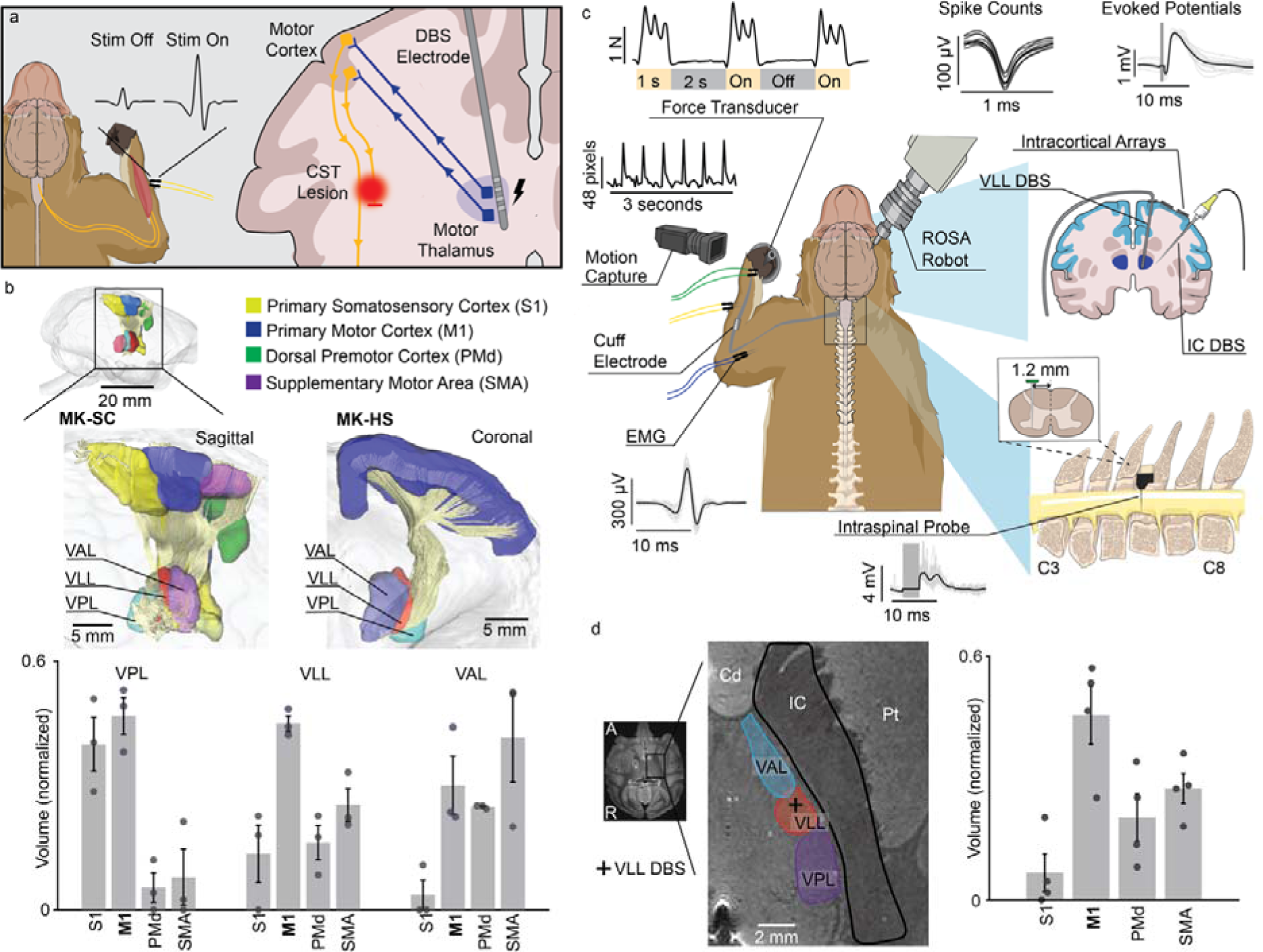
Identification of optimal target of stimulation. **(a)** Schema of our hypothesis: motor thalamus stimulation potentiates motor output by increasing excitability of the motor cortex. The potentiation persists also after a lesion of the CST highlighted in red. **(b)** *Top*: High definition fiber tracking (HDFT) from VPL, VLL, and VAL nuclei (VPL: ventral posterolateral, VLL: ventral laterolateral, VAL: ventral anterolateral) of monkey motor thalamus to cortical regions (n=3) (S1: primary somatosensory cortex, M1: primary motor cortex, PMd: dorsal pre-motor cortex, SMA: supplementary motor area). *Bottom*: Volume of thalamocortical projections (mean ± standard error over 3 animals) from each nucleus to each cortical region normalized by the total volume of fibers projecting from each nucleus. **(c)** Acute experimental setup. First, a cuff electrode was implanted around the motor branch of the radial nerve for stimulation. Animals were then implanted with a DBS electrode in the internal capsule (IC) and one in the VLL using the ROSA robot and intracortical arrays over S1 and M1. An intraspinal probe was implanted at the C6 spinal segment to record spinal local field potentials and EMG needle electrodes were inserted in arm, hand, and face muscles. A force transducer was placed in the animal’s hand to measure grip force. Finally, a camera recorded the kinematic of the arm and hand. **(d)** *Left*: Example of VLL electrode implant location localized from post-mortem MRI (Cd: Caudate Nucleus, IC: Internal Capsule, Pt: Putamen). *Right*: Normalized volume HDFT projections from the area of stimulation to cortical regions (mean ± SE over animals, n=4).

### Targeted deep brain stimulation of the VLL specifically excites the motor cortex

Previous studies have revealed that the thalamocortical projections, as identified with our HDFT analysis, are mostly glutamatergic^12,24^. We therefore hypothesized that stimulation of the VLL would excite the motor cortex. To demonstrate this hypothesis, we implanted a human-grade electrode (Microdeep® SEEG Electrodes, DIXI Medical, Marchaux-Chaudefontaine, France) within the VLL nucleus in anesthetized monkeys (n=5) using the ROSA ONE® Robot Assistance Platform (ROSA robot, Zimmer Biomet, Warsaw, Indiana, USA; **Fig. 1c** and **Extended Data Fig. 1**). High-resolution post-mortem structural MRI confirmed the accurate location of the DBS electrode within the VLL (average distance between the implantation location and the VLL nucleus over animals:1.7mm +/− 0.4 mm (SE), in line with human studies^25^). Additionally, we used HDFT between the electrode implantation region and the somatosensory, motor, and pre-motor cortical areas and confirmed that the largest volume of fibers within the stimulation area projected to M1 (on average 55% of the fibers) (**Fig. 1d**).

In addition to the DBS electrode within the VLL nucleus, we implanted two 48-channels intracortical arrays (Utah arrays) in the primary motor (M1) and somatosensory (S1) cortices, respectively (**Fig. 1c**). In n=4 anesthetized monkeys, we applied continuous stimulation of the VLL at 10 Hz (see **Supplementary Table 2** for details on stimulation parameters) while recording local field potentials and multi-unit neuronal activity from these cortical areas (**Fig. 2a**). We then analyzed VLL stimulation triggered averages of evoked potentials (EPs) in M1 and S1 electrodes and observed clear cortical EPs with peak latencies at 5-10 ms post-stimulation (see example **Fig. 2b**) confirming a direct monosynaptic pathway between the VLL and cortex^26,27^. Analysis of EPs peak to peak amplitude across the entire arrays showed significantly larger responses in M1 as compared to S1 in all monkeys (see histograms **Fig. 2b**), confirming preferential projections of VLL axons towards M1. As expected, these responses were larger along the pre-central gyrus where the hand and arm representation in M1 are located (see heat maps **Fig. 2b**). Additionally, stimulation-evoked spike counts corroborated these findings showing an increase in multi- and single-units firing rates post-stimulation (**Fig. 2c**), which was significantly higher in M1 as compared to S1 in both monkeys.

**Fig. 2:**
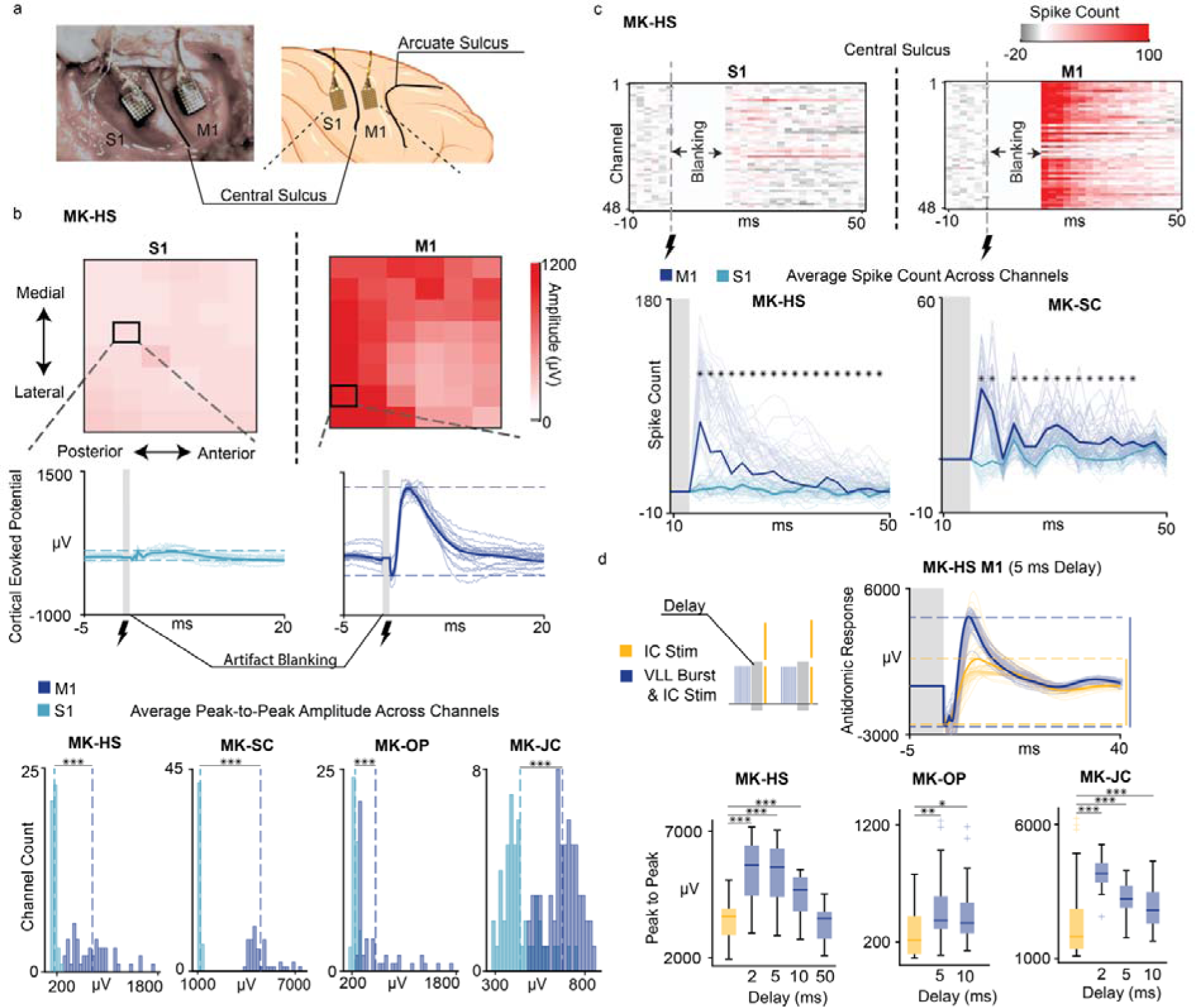
Stimulation of the motor thalamus increases motor cortex excitability. **(a)** Picture (*left*) and schematic (*right*) of the Implant location of M1 and S1 intracortical arrays. **(b)** *Top*: Example heatmap of average peak to peak amplitudes of cortical evoked potentials from VLL stimulation at 10 Hz across all channels over S1 and M1 for MK-HS. *Center*: Example stimulation triggered averages of cortical evoked potentials over S1 and M1 (n=40 traces). *Bottom*: Histogram of peak to peak amplitudes across all channels for S1 and M1 (n=48 channels per array). **(c)** Top: Example baseline corrected spike count heatmaps in S1 and M1 for MK-HS. *Bottom*: Average spike counts over time across all channels in S1 and M1 array (n=48 channels per array). **(d)** *Top*: Example traces of antidromic potentials in M1 from IC stimulation without (yellow) and with (blue) conditioning from a burst of VLL stimulation for MK-HS (n=40 traces). Boxplot of peak to peak amplitude of the antidromic potentials when IC stimulation is conditioned by VLL stimulation at various delays (2, 5, 10, and 50ms). In the boxplot, the whiskers extend to the maximum spread not considering outliers, central, top, and bottom lines represent median, 25^th^, and 75^th^ percentile, respectively. For all panels, statistical significance was assessed with two-tail bootstrapping with Bonferroni correction: p<0.05 (*), p<0.01 (**), p<0.001(***).

Cortical EPs elicited from VLL stimulation should be indicative of increased excitation of cortico-spinal neurons located in M1. If this is true, the amplitude of antidromic neural responses elicited in cortical-spinal axons from stimulation of the CST and recorded in M1 should be larger when VLL stimulation is active, because of decreased cortical thresholds in these neurons. To test this hypothesis, a stimulating electrode was implanted into the posterior limb of the internal capsule (IC, **Fig. 1c**), which contains the cortical-spinal axons originating in the motor cortex and projecting to the spinal cord. We targeted the hand representation of the CST. Direct stimulation of these axons via single pulse stimulation to the IC elicited antidromic action potentials toward the cell body of the pyramidal neurons in M1 at short latencies (5ms post-stimulation) (**Fig. 2d**). We then conditioned the single pulse stimulation to the IC by a 100ms burst at 100 Hz to the VLL nucleus at different delays (2-50ms). We found that the peak to peak amplitude of the antidromic neural responses in M1 was significantly higher when a burst of VLL stimulation preceded IC stimulation (n=3) (**Fig. 2d**) confirming that VLL stimulation increases excitability of cortico-spinal neurons in M1. At delays longer than 10ms, the antidromic potentials returned to amplitudes similar to those of IC stimulation alone.

Overall, these results demonstrate that stimulation of the VLL nucleus increases the excitability of cortico-spinal neurons specifically within M1.

### Motor thalamus stimulation potentiates upper-limb motor outputs

We built on previous experiments that reported modulated MEPs from TMS of the motor cortex when conditioned with stimulation of VIM, GPi, or STN^7–10^, and we hypothesized that the enhanced cortical excitability observed in our monkeys would lead to amplified MEPs recorded in arm, hand, and finger muscles. To test this, we paired stimulation of the IC at 2 Hz with continuous stimulation of VLL at 50 or 80 Hz (**Fig. 3a**), i.e., frequencies often used in other neuromodulation applications such as spinal cord stimulation^28^. Stimulation of the IC at 2 Hz generated short latency (on average 10-15 ms) MEP responses in hand, arm, and finger muscles (**Fig. 3b**) as well as movements of the limb (**Extended Data Fig. 3**) suggesting a monosynaptic activation of spinal motoneurons. When 2 Hz IC stimulation was paired with continuous stimulation of the VLL at 50 and 80 Hz, the area under the curve (AUC) of MEPs and the peak to peak amplitude of movements of the arm (n=2), hand (n=3), and fingers (n=3) were immediately and significantly increased (p < 0.001, **Fig. 3b**, **Extended Data Fig. 2** and **3**, **Video 1**). Interestingly, a dose response relationship was visible as MEP amplitudes increased proportionally when increasing the VLL stimulation amplitude (**Fig. 3a**). Additionally, MEPs of face muscles were also significantly larger with stimulation of the VLL at 50 and 80 Hz (**Extended Data Fig. 2a**).

**Fig. 3:**
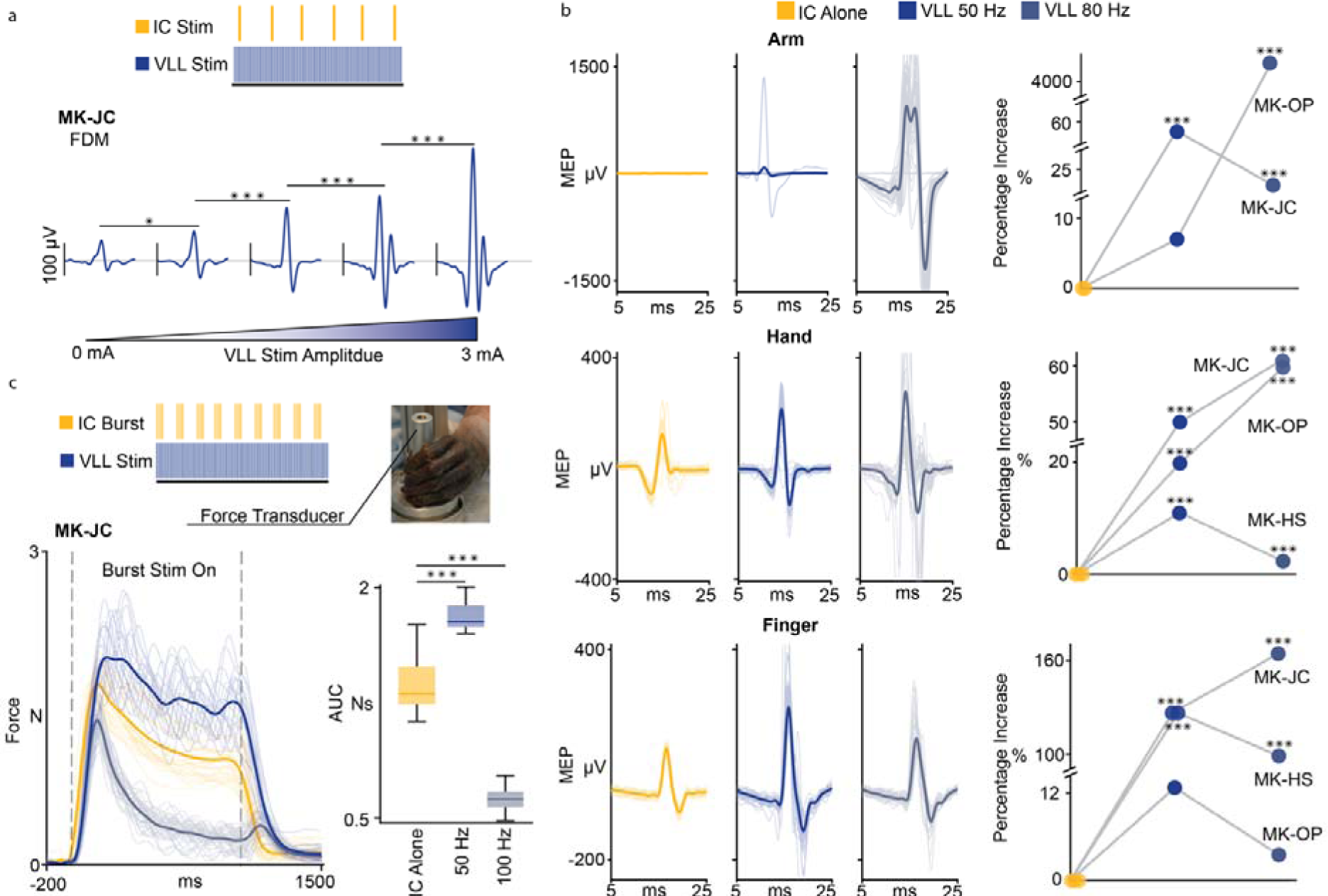
Stimulation of the motor thalamus amplifies motor outputs. **(a)** Average of binned Flexor Digitorum Minimi (FDM) MEPs generated by IC stimulation at 2 Hz paired with continuous VLL stimulation at 50 Hz with gradual ramp up of amplitude (0 to 3mA; bins: 0 to 0.6 mA, 0.7 to 1.2 mA, 1.3 to 1.8 mA, 1.9 to 2.4 mA, and 2.5 to 3 mA, each bin included n=9 responses). **(b)** *Left panels*: MEPs of one arm muscle (n=40, Biceps, Mk-OP), one hand muscle (n=40, Extensor Digitorum Communis, Mk-HS), and one finger muscle (n=40, Abductor Pollicus Brevis, MK-HS) with IC stim alone and then paired with VLL stimulation at 50 and 80 Hz. *Right panels*: percentage of increase of AUC of arm, hand, and finger MEPs between IC stimulation alone and paired with VLL stimulation at 50 and 80 Hz. For each monkey, the percentage of increase was calculated over the medians and averaged over all the muscles. See **Extended Data Fig. 2** for box-plots for single muscles. (***), (**), or (*) was placed if muscles in each group show a significant increase (respective to p-values 0.001, 0.01, and 0.05). **(c)** Force transducer experimental setup and stimulation parameters (IC: 45-50 Hz burst, 1s ON, 2s OFF; VLL: 50 Hz continuous). Example force traces (n=20) and boxplots of AUC (IC alone, IC with VLL at 50 Hz, and IC with VLL at 100 Hz). For all boxplots, the whiskers extend to the maximum spread not considering outliers, central, top, and bottom lines represent median, 25^th^, and 75^th^ percentile, respectively. For all panels, statistical significance was assessed with two-tail bootstrapping with Bonferroni correction: p<0.05 (*), p<0.01 (**), p<0.001(***).

Finally, in order to demonstrate that VLL stimulation could enhance induced functional movements, we delivered bursts of IC stimulation at about 50 Hz which induced a grasping-like motion producing measurable isometric forces (**Fig. 3c**). We then paired the functional IC stimulation with VLL stimulation at 50 or 100 Hz. VLL stimulation at 50 Hz, but not at 100 Hz, immediately and significantly increased the grip force as compared to IC stimulation alone (n=1) (**Fig. 3c**, **Video 2**).

In summary, targeted motor thalamus stimulation increased upper-limb motor output as measured by the amplitude of arm and hand muscle MEPs and movement kinematics, and stimulation-induced grip forces.

### Control experiments: motor output potentiation occurs through orthodromic thalamocortical pathways

We sought to confirm that the enhanced motor outputs was the result of an increased excitability of the motor cortex and did not result instead from inadvertent activation of descending excitatory tracts due to current spreading from motor thalamus stimulation. We reasoned that if the observed MEP potentiation was occurring within the spinal cord as a result of inadvertent stimulation of descending tracts, then we would expect that peripheral inputs to spinal circuits, such as H-reflexes, would also be potentiated by VLL stimulation. To control for this, we implanted a multi-channel linear probe (NeuroNexus) at the C6-C7 spinal level with dorsal-ventral orientation (**Fig. 1c**). We observed consistent volleys in the spinal cord in the first 3 ms following VLL stimulation (1.44 ms, 1.73, and 2.13 ms for Mk-OP, Mk-JC, and Mk-HS, respectively, **Fig. 4a**) with largest peak to peak amplitude in the intermediate-ventral spinal cord gray zone (**Fig. 4b**). These evoked potentials could either represent antidromic recruitment of ascending pathways such as spino-thalamic axons, or orthodromic activation of descending excitatory pathways. However, there were no MEPs in either the upper-limb or facial muscles when stimulating the VLL nucleus alone (**Fig. 4c**, **Extended Data Fig. 4**) suggesting that the observed spinal responses were not caused by inadvertent stimulation of the CST. Moreover, when we simultaneously stimulated the VLL and the IC, the frequency of the muscle contractions was at 2 Hz (i.e., same frequency of IC stimulation) and not at the frequencies of VLL stimulation (50, 80, or 100 Hz) (**Extended Data Fig. 3**), further demonstrating that the increase in MEPs and movement kinematic were not induced by current spreading to the CST from the VLL. It was still possible that the observed spinal responses and the enhanced movements could be carried by other descending tracts that do not have direct spinal motor neuron connections or generate MEPs but that are able to excite the spinal circuits and facilitate movement. However, when we paired continuous VLL stimulation at 50 Hz with stimulation of the radial nerve we observed no significant increase of reflex-mediated responses (**Fig. 4d** and **Extended Data Fig. 5**). These results demonstrate that 1) the observed volleys in the spine are likely the result of antidromic activation of ascending pathways such as spino-thalamic axons; and 2) the observed potentiation of MEPs (**Fig. 3**) is not occurring inside the spinal cord as a result of current spread to descending axons, but is, instead, a trans-cortically mediated effect.

**Fig. 4:**
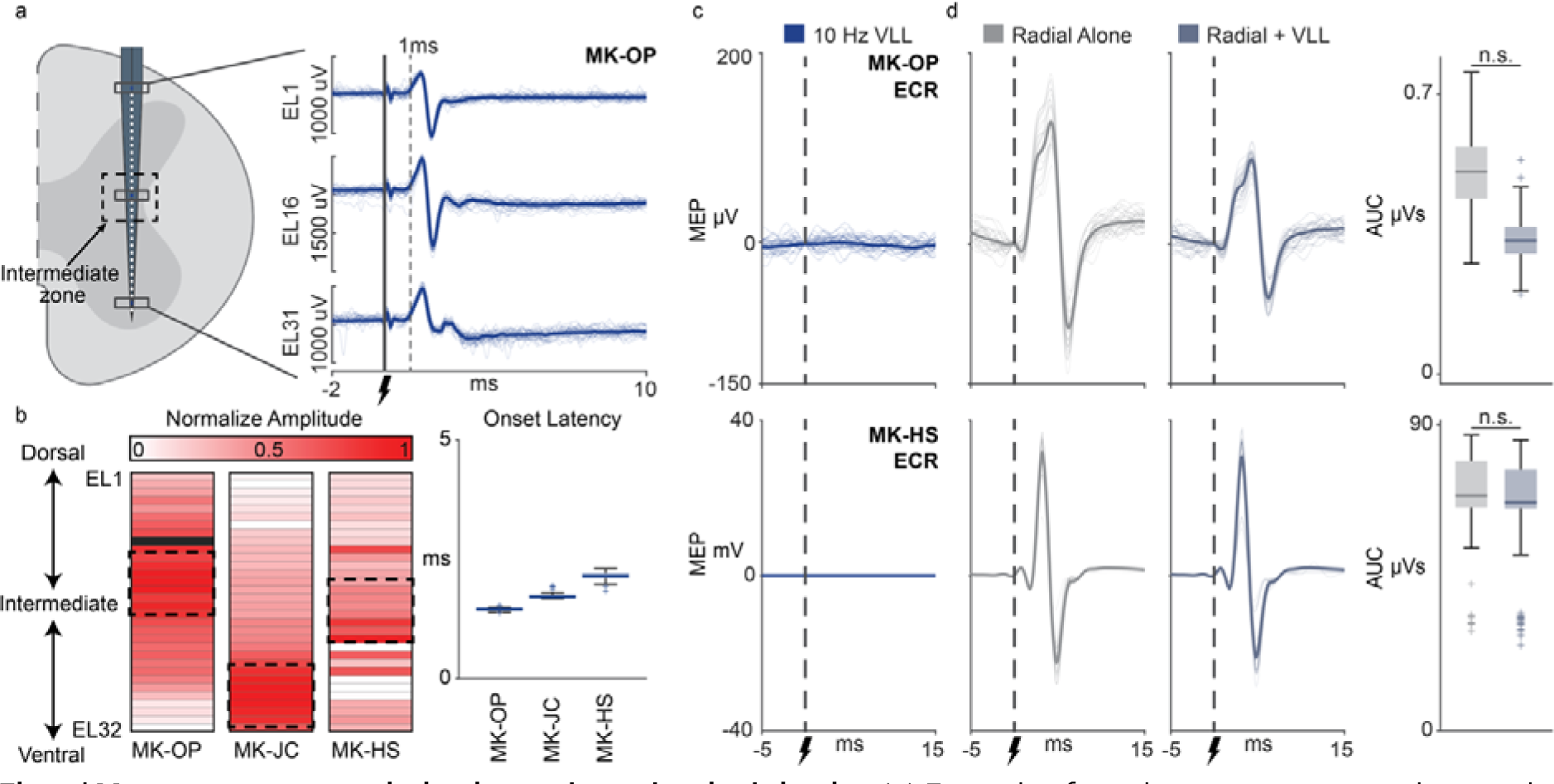
Motor output potentiation is not through spinal circuits. **(a)** Example of antidromic responses in the spinal cord from VLL stimulation at 10 Hz (n=30 traces) in MK-OP. **(b)** *Left*: Heatmaps of peak to peak amplitude of the antidromic responses in the spinal cord at the C6-C7 spinal level with dorsal-ventral alignments for n=3 animals. The dashed boxes are highlighting the putative intermediate-ventral zone where we see the greatest responses. *Right*: Boxplots of the antidromic response latency for each animal (n = 656 MK-OP, n = 596 MK-JC, n = 289 MK-HS). **(c)** MEPs of the wrist (ECR: Extensor Carpi Radialis, 30 traces for each animal) elicited by VLL stim at 10 Hz. **(d)** EMG reflexes and boxplots of the AUC of the EMG reflexes of the ECR muscle elicited by radial nerve stimulation and radial nerve paired with continuous VLL stimulation at 50 Hz (30 example traces each). For all boxplots, the whiskers extend to the maximum spread not considering outliers, central, top, and bottom lines represent median, 25^th^, and 75^th^ percentile, respectively. Statistical significance was assessed with one-tail bootstrapping with Bonferroni correction, however, in all cases the results were not significant.

### Efficacy of VLL stimulation in promoting motor augmentation is frequency dependent

Our results suggest that the potentiation of descending CST activity is mediated by the orthodromic recruitment of cortico-spinal neurons in M1 via thalamocortical excitatory synaptic projections. Excitatory synaptic transmission is known to be affected by stimulation frequency via mechanisms such as presynaptic inhibition or homosynaptic neurotransmitter suppression^29^. Accordingly, we explored the effect of multiple frequencies on the efficacy of motor potentiation. We delivered IC stimulation at 2 Hz paired with continuous VLL stimulation at 10, 50, 80, 100, and 200⍰Hz. We analyzed the MEP responses to the individual IC stimulation pulses over time and observed a variety of modulation patterns (**Fig. 5a**). We classified these patterns into four distinct categories: (**1**) *no potentiation*, as responses were comparable to IC stimulation alone, (**2**) *sustained potentiation*, as responses were potentiated, (**3**) *attenuation*, as the response amplitudes progressively decreased compared to the initial responses, and (**4**) *suppression*, as the responses were completely suppressed after a few pulses. Given that both *attenuation* and *suppression* result in decreased MEP responses over time, we combined them in **Fig. 5b** (see **Extended Data Fig. 2d** for the separated *attenuation* and *suppression*). Generally, the absence of potentiation of the IC stimulation responses was characteristic of low frequencies (10 Hz, observed in > 50% of the muscles across all 3 animals), whereas stimulation frequencies between 50 Hz and 80 Hz consistently potentiated the MEP amplitudes with sustained outputs (**Fig. 5b**). Importantly, attenuation/suppression was only present in less than 15% of the muscles for 50 Hz and less than 25% of the muscles for 80 Hz. Stimulation at 100 Hz and above, instead, resulted mainly in attenuation and suppression and only in a few instances of sustained potentiation. Therefore, we identified the 50-80 Hz range as optimal to achieve sustained potentiation of MEPs.

**Fig. 5:**
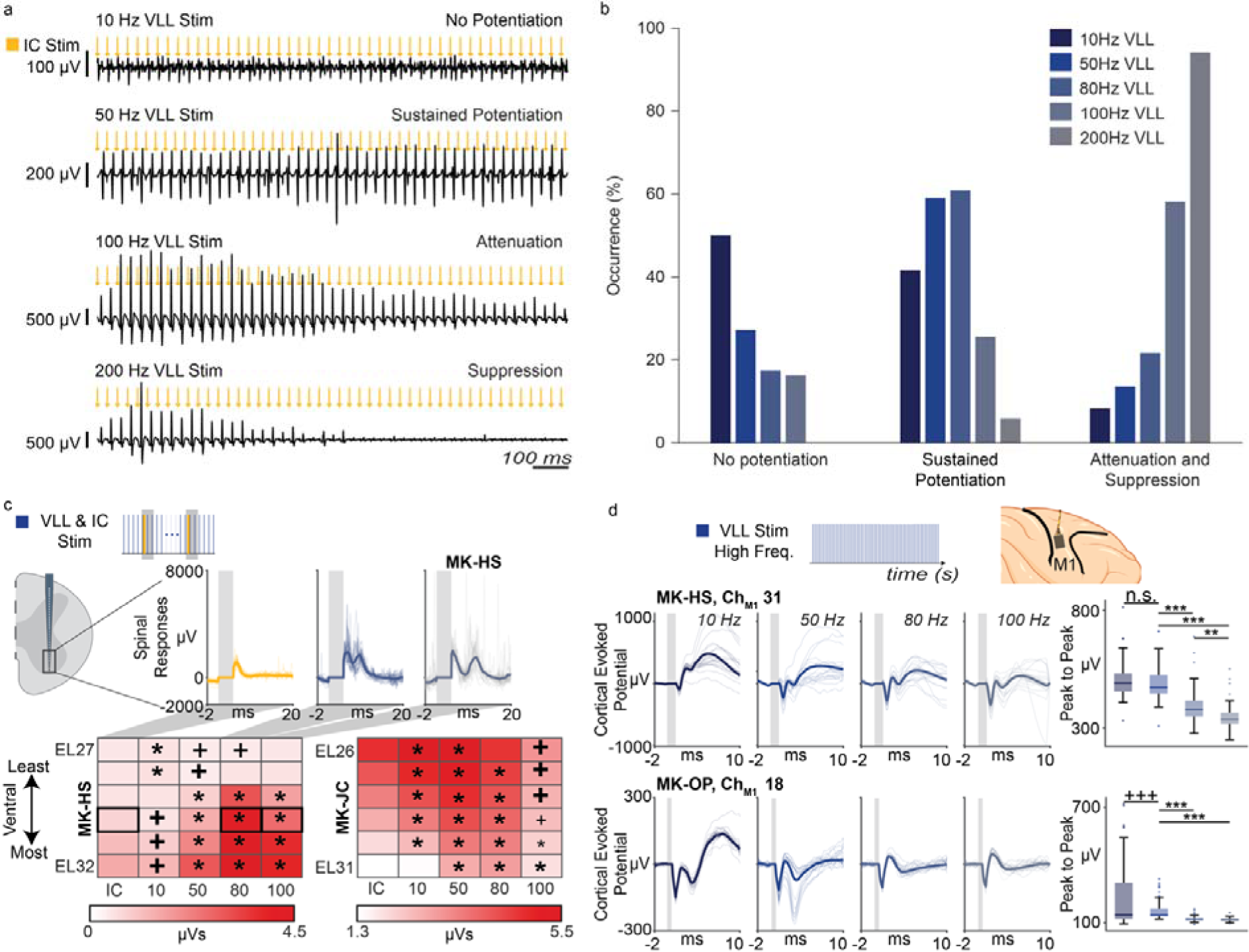
Responses are modulated in a frequency-dependent manner. **(a)** Examples of frequency-dependent modulation of muscular responses. EMG responses were elicited by 2 Hz stimulation of the IC paired with different VLL stimulation frequencies (10, 50, 80, 100, and 200 Hz). **(b)** The occurrence of modulation patterns with respect to stimulation frequency. All patterns recorded in all muscles of 3 animals were included in the analysis (n=E24 patterns at 10⍰Hz, n=22 patterns at 50⍰Hz, n=23 patterns at 80⍰Hz, n=43 at 100 Hz, and n=17 patterns at 200⍰Hz). **(c)** *Top*: Example of spinal responses in the ventral zone for IC stimulation alone and IC stimulation paired with VLL stimulation at 80 and 100 Hz (n = 30 traces per plot). *Bottom*: Heatmaps of the AUC calculated from 5 to 10 ms after IC stimulation for all ventral channels for IC stimulation alone and IC stimulation paired with VLL stimulation at 10, 50, 80, and 100 Hz. (*) for significant potentiation and (+) for significant suppression. Bold represents p<0.001. **(d)** *Top*: Schematic of the experimental layout for testing frequency dependence within the motor cortex. *Bottom*: Example traces of the cortical evoked potential responses in the M1 array when stimulating the thalamus at different frequencies (10, 50, 80, and 100 Hz) (n = 30 traces). Boxplots of the peak to peak amplitudes of the cortical evoked potentials. Statistical significance was tested by comparing 50 Hz VLL stimulation to all other stimulation conditions for potentiation using one-tailed bootstrapping with Bonferroni correction: p<0.05 (*), p<0.01 (**), p<0.001(***). For all boxplots, the whiskers extend to the maximum spread not considering outliers, central, top, and bottom lines represent median, 25^th^, and 75^th^ percentile, respectively.

We further explored the neural correlates of these effects with intra-cortical and intra-spinal neural recordings. IC stimulation pulses elicited intra-spinal neural responses in the ventral zone^30^, where spinal motoneurons are located. Consistent with the EMG recordings, the AUC of these responses were significantly larger when the VLL nucleus was stimulated at 50 or 80 Hz (p < 0.05, **Fig. 5c**). Spinal responses were, instead, suppressed at higher frequencies. Similarly, cortical EPs in M1 when stimulating the VLL alone showed larger peak to peak amplitudes with frequencies of stimulation in the 50-80 Hz range as compared to 100 Hz (**Fig. 5d**). These combined results demonstrate that the effects observed in the MEP and spinal cord responses were a consequence of frequency dependent excitation of the motor cortex. This frequency dependent characteristic of excitation could be either the result of presynaptic inhibition or homosynaptic neurotransmitter suppression at the level of the cortico-spinal neurons^18,28,29^.

In summary, our analysis confirmed a frequency dependent effect that is consistent with motor cortical potentiation of thalamic afferents to M1 cortex via excitatory synaptic inputs. From these findings, we identified that the optimal stimulation frequencies to increase muscle outputs are in the 50-80 Hz range, notably lower than clinical DBS stimulation frequencies (> 100 Hz)^31^.

### Potentiation of motor outputs persists after CST lesions

To demonstrate the potential clinical relevance of cortical potentiation, we examined whether electrical stimulation of the VLL nucleus would still augment motor outputs after a partial lesion of the CST. Such a lesion reduces the number of CST fibers projecting to the cervical spinal cord, leading to motor deficits similar to those observed in human patients. To create such a lesion, we implanted a second depth electrode in the posterior limb of the IC, approximately 2 cm ventral/inferior to the IC stimulating electrode. We then generated a controlled and reproducible lesion of the CST fibers by performing thermo-frequency ablations^32^ through the ventral/inferior electrode (**Fig. 6a**). As expected, we observed reduced MEP amplitudes when stimulating the dorsal IC electrode after the ventral/inferior CST lesion had been made (**Extended Data Fig. 6**). Post-mortem HDFT confirmed that the lesion consistently and significantly reduced the volume of CST fibers compared to the intact hemisphere in all tested monkeys (46% reduction) (n=4) (**Fig. 6a**). After the CST lesion, the smaller MEPs recorded from the arm, hand and fingers muscles were significantly increased when the VLL nucleus was simultaneously stimulated at 50 and 80 Hz (p < 0.001, **Fig. 6b** and **Extended Data Fig. 7b**). Importantly, IC and VLL stimulation intensity were the same of those used pre-lesion. MEP amplitudes when VLL stimulation was ON were comparable to pre-lesion MEP amplitudes in the same muscles (**Extended Data Fig. 6**). Furthermore, post-lesion stimulation at 80 Hz resulted in a stronger MEP potentiation as compared to 50 Hz. Similarly, the grip force elicited by IC stimulation bursts significantly increased with continuous VLL stimulation at 50 Hz and an even greater increase occurred with VLL stimulation at 80 Hz (**Fig. 6d**, **Video 3**). Finally, face muscles MEPs were also potentiated by VLL stimulation after the CST lesion (**Extended Data Fig. 6**).

**Fig. 6:**
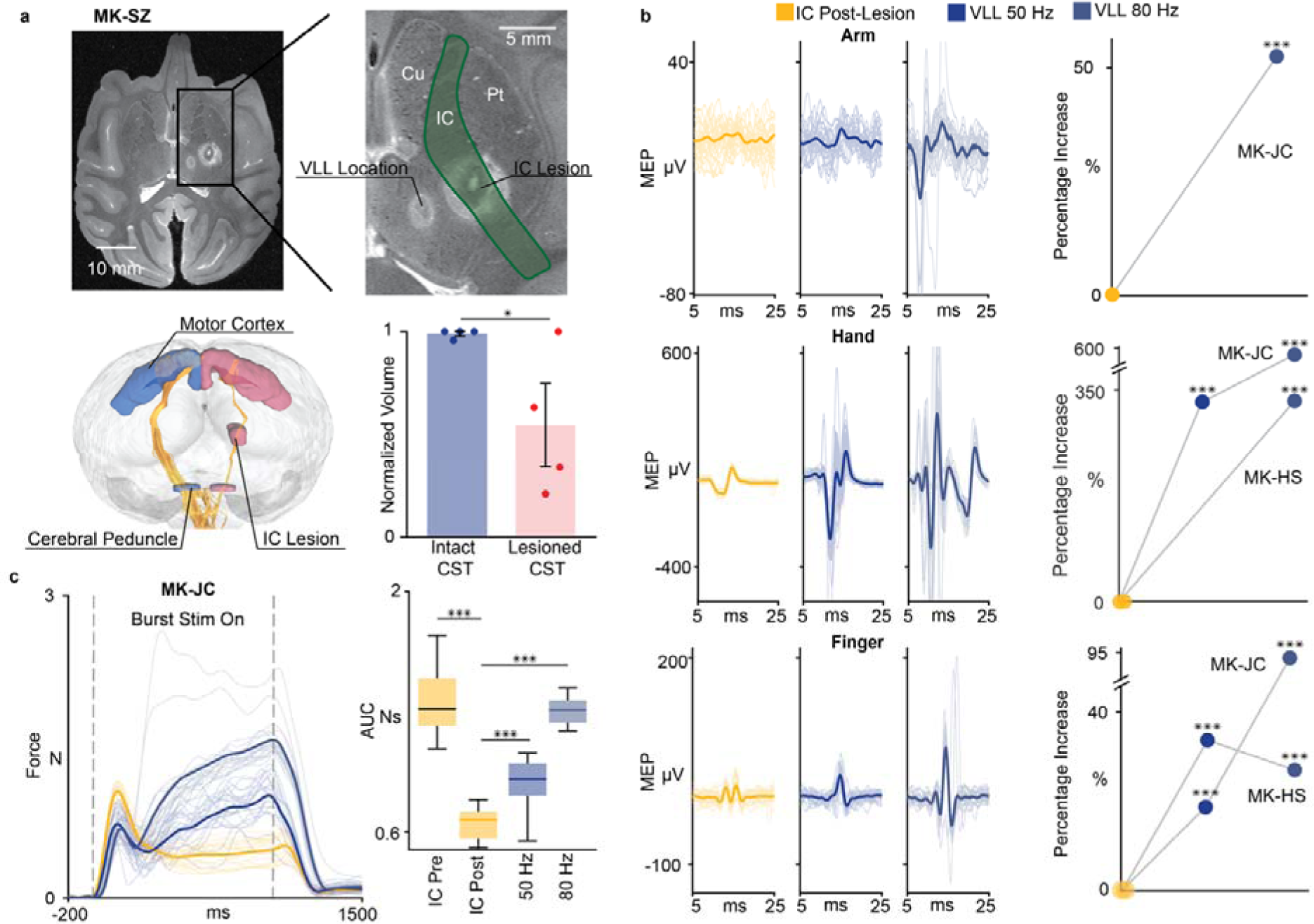
Stimulation of the motor thalamus amplifies motor outputs after CST lesions. **(a)** *Top panels*: T2-weighted post-mortem MRI of IC lesion and VLL location (axial plane). (Cu: Caudate Nucleus, IC: Internal Capsule, Pt: Putamen). *Bottom panels*: HDFT of the CST in intact and lesioned hemispheres. Volume of cumulative CST (mean +/− SE over animals) for both hemispheres normalized over the volume of the intact hemisphere. **(b)** *Left panels*: Example of post-lesion MEPs of one arm muscle (n=40, BIC: Biceps, MK-JC), one hand muscle (n=40, ECR: Extensor Communis Radialis, Mk-JC), and one finger muscle (n = 40, FDM: Flexor Digitorum Minimi, MK-JC) with IC stim alone and then paired with VLL stimulation at 50 and 80 Hz. *Right panels*: percentage of increase of arm, hand, and finger MEPs post-lesion between IC stimulation alone and paired with VLL stimulation at 50 and 80 Hz. For each monkey, the percentage of increase was calculated over the medians and averaged over all the muscles. See **Extended Data Fig. 6** for box-plots for single muscles. (***), (**), or (*) was placed if muscles in each group show a significant increase (respective to p-values 0.001, 0.01, and 0.05). **(c)** *Left panel*: Example of force traces (n=20). *Right panel*: boxplot of AUC pre- and post-lesion for IC alone, and IC with VLL 50 Hz and VLL 80 Hz. For all boxplots, the whiskers extend to the maximum spread not considering outliers, central, top, and bottom lines represent median, 25^th^, and 75^th^ percentile, respectively. For all panels, statistical significance was assessed with two-tail bootstrapping with Bonferroni correction: p<0.05 (*), p<0.01 (**), p<0.001(***).

Overall, these results demonstrate that VLL stimulation can potentiate motor output even in the presence of hemiparesis caused by partial lesions of the CST.

### Stimulation of the motor thalamus potentiates motor output in humans

We sought to verify the translational potential of these results in human subjects. For this, after obtaining informed written consent, we performed intraoperative electrophysiological experiments in human subjects (n=4, 2 males, 2 females) who underwent on-label DBS implants in the motor thalamus (**Fig. 7a**). Experiments were designed to closely mimic the animal study protocols. The human motor thalamus nuclei comparable to the VLL nucleus are the ventralis intermediate (VIM) and the ventralis oralis posterior (VOP). HDFT analysis confirmed preferential anatomical projections from the VIM/VOP to M1 (**Fig. 7b**). Additionally, we tested the connectivity of these pathways using evoked potential methods. For this, we placed a 6-channel subdural strip electrode (Adtech, Oak Creek, WI, USA) over the arm and hand representation of the primary motor and somatosensory cortices. To confirm electrode placement, we used the validated clinical technique of somatosensory evoked potential (SSEP) phase reversal (PR) mapping^33^ to locate both the hand representation of S1 (largest amplitude N20/P30 cortical SSEP) and the approximate location of the central sulcus, where the polarity of the SSEP reverses (**Fig. 7a**). We then mapped the upper extremity representation of M1 within the precentral gyrus by direct cortical stimulation (DCS) of the electrode contacts that were anterior to the SSEP PR. MEPs were recorded from six contralateral upper extremity muscle groups (Deltoid, Biceps, Triceps, Flexor carpi, Extensor carpi and APB). The strip electrode position was adjusted to obtain MEPs from at least three muscle groups with the lowest possible stimulation threshold (∼6-12 mA).

**Fig. 7:**
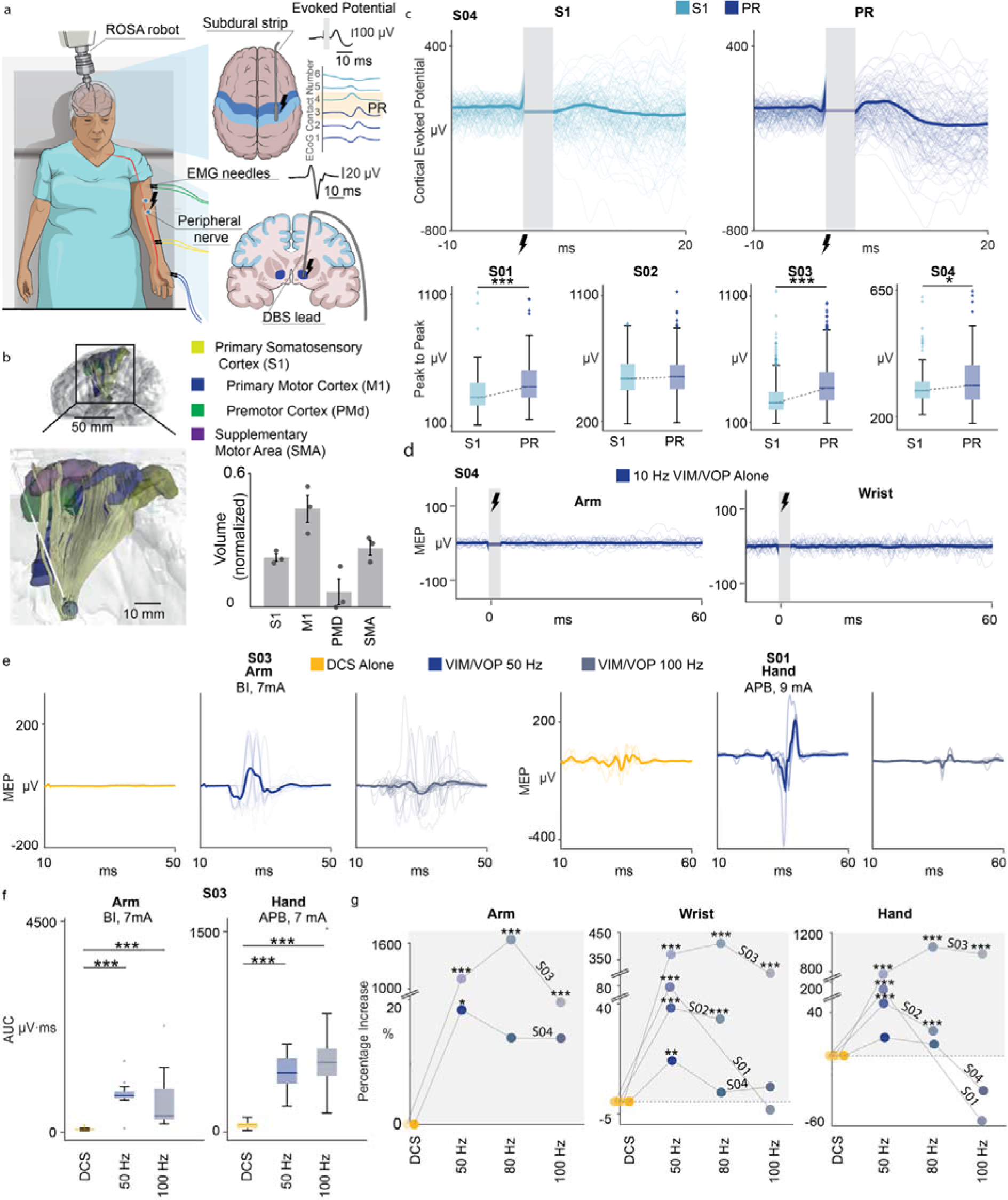
Stimulation of the motor thalamus amplifies motor outputs in humans. **(a)** *Top*: Experimental setup for human intraoperative experiments. Enlargement shows a schematic representing the subdural strip electrode placement over the primary motor (M1) and somatosensory (S1) cortices, and the phase reversal (PR) to identify the central sulcus. Needle electrodes were inserted in arm, wrist, and hand muscles to record MEPs and superficial electrodes were placed over the median nerve for SSEP. **(b)** *Left*: HDFT from the VIM/VOP to cortical regions. *Right*: Normalized volume (mean ± SE over n=4 subjects) of VIM/VOP projections to each cortical region normalized by the total volume of fibers. **(c)** *Top*: Example traces (n=122) of cortical evoked potentials elicited by VIM/VOP stimulation recorded over an S1 (left) and PR (right) contact for S04. *Bottom*: Box-plots of peak to peak amplitude of cortical evoked potentials at S1 and PR contact. From left to right, subjects 1 to 4 are shown (n=128, n=585, n=601, n=122 trials, respectively). **(d)** Example MEP traces (arm, biceps; hand, flexor; n=60) with VIM/VOP stimulation alone for S03. **(e)** Example MEP traces with DCS alone and DCS paired with VIM/VOP stimulation at 50 and 100 Hz (from left to right). Arm is S03 biceps (n=48 traces), fingers is S01 abductor pollicis brevis (n=16 traces). **(f)** Box plots of AUC for MEPs of the arm and fingers (biceps and abductor pollicis brevis respectively; n=58) with DCS alone and DCS paired with VIM/VOP stimulation at 50 and 100 Hz. **(g)** Scatter plots for arm, hand, and fingers muscles of all subjects, representing the percentage of AUC increase calculated over the means, with respect to DCS alone, for all the different VIM/VOP stimulation frequencies (50, 80, and 100 Hz). For all boxplots, the whiskers extend to the maximum spread not considering outliers, central, top, and bottom lines represent median, 25^th^, and 75^th^ percentile, respectively. For all panels, statistical significance was assessed with two-tail bootstrapping with Bonferroni correction: p<0.05 (*), p<0.01 (**), p<0.001(***).

We then examined the projections from VIM/VOP to S1 and M1 upper extremity representations by recording cortical EPs at each of the 6 electrode contacts in response to low frequency (2 or 10 Hz) stimulation of the VIM/VOP DBS electrode (Boston Scientific) implanted in the same hemisphere at the AC/PC plane (X: 12mm lateral to the AC/PC line, Y: 6mm anterior to PC, Z: 0mm to the AC/PC horizontal plane, **Extended Data Fig. 8a**). Cortical EPs, occurring within 20 ms of the stimulus onset, were recorded at the M1 contacts (anterior to the PR location) (**Fig. 7c**). In all subjects but one, the peak to peak amplitude of these evoked responses was significantly larger for precentral contacts than for postcentral contacts. This supports orthodromic synaptic transmission through thalamocortical projections from VIM/VOP to the M1 hand/arm representation as was observed in monkeys during stimulation of the VLL.

Next, DCS MEPs generated from stimulating the optimal contact over M1 cortex were recorded from up to six contralateral upper extremity muscles without and with paired VIM/VOP stimulation at 50, 80, or 100 Hz. Similar to the monkey experiments, we observed a consistent and statistically significant increase in the DCS MEP amplitudes across arm, hand, and fingers muscles with VIM/VOP stimulation at 50-80 Hz compared to DCS alone (p < 0.001, **Fig. 7f**). A more variable behavior, instead, was found with VIM/VOP stimulation at higher frequencies. Indeed, stimulation at 100 Hz further amplified or suppressed these responses (**Fig. 7g**, **Extended Data Fig. 9**). Interestingly, suppression was predominant for the finger muscles similar to the monkeys (**Fig. 3b, Extended Data Fig. 2**). Finally, we confirmed that electrical stimulation of the VIM/VOP alone did not produce motor evoked potentials in the arm, hand, nor fingers muscles (**Fig. 7d**), similar to the monkeys’ experiments.

Overall, these results observed in human subjects were equivalent to those obtained in monkeys, demonstrating the translational potential of the mechanisms of action and of the identified targets and stimulation parameters. Specifically, electrical stimulation of the motor thalamus enhanced motor cortex excitability at specific stimulation frequencies and consequently potentiated motor output via the CST in human subjects.

### Stimulation of the motor thalamus increases motor output in a person with CST lesion

We had the opportunity to test our hypothesis in one patient who suffered severe bilateral lesions of the CST (CST01, **Fig. 8a**) and underwent on-label DBS implants in the motor thalamus (**Extended Data Fig., 8b**). We quantified damage to the white-matter axons using differential tractography and verified that for both hemispheres the most damaged tract was the CST (57% of damaged tracts in the right hemisphere and 89% in the left hemisphere). Consequently, CST01 suffered from hemiparesis of both upper extremities. Functionally, CST01 required maximal assistance for eating, grooming, bathing, and dressing. Replicating the same human set-up (**Fig. 7a**), a 6-contact subdural strip electrode was placed over the S1 and M1 cortex of the left hemisphere (i.e., greater CST damage). Consistent with the results in monkeys and human subjects, stimulation of the VIM/VOP nucleus resulted in larger amplitude cortical EPs over motor cortical areas as compared to somatosensory cortex (**Fig. 8b**). Additionally, DCS MEPs were consistently recorded from the APB and flexor muscles of the hand, even if, because of the lesion, MEPs were smaller for the finger muscles (**Fig. 8c**, left panels). We then paired DCS of M1 with stimulation of the motor thalamus to assess the effect of VIM/VOP stimulation on the hemiparetic arm. Again, DCS MEPs were significantly larger with concurrent stimulation of the motor thalamus at 50 Hz and modulated in a frequency-dependent manner (**Fig. 8c**, right panels). Indeed, stimulation at 100 Hz resulted in suppression of MEPs particularly for the fingers muscles, demonstrating the consistency of our findings across multiple monkeys and human subjects, including one patient suffering from a chronic (> 6 months) CST lesion.

**Fig. 8:**
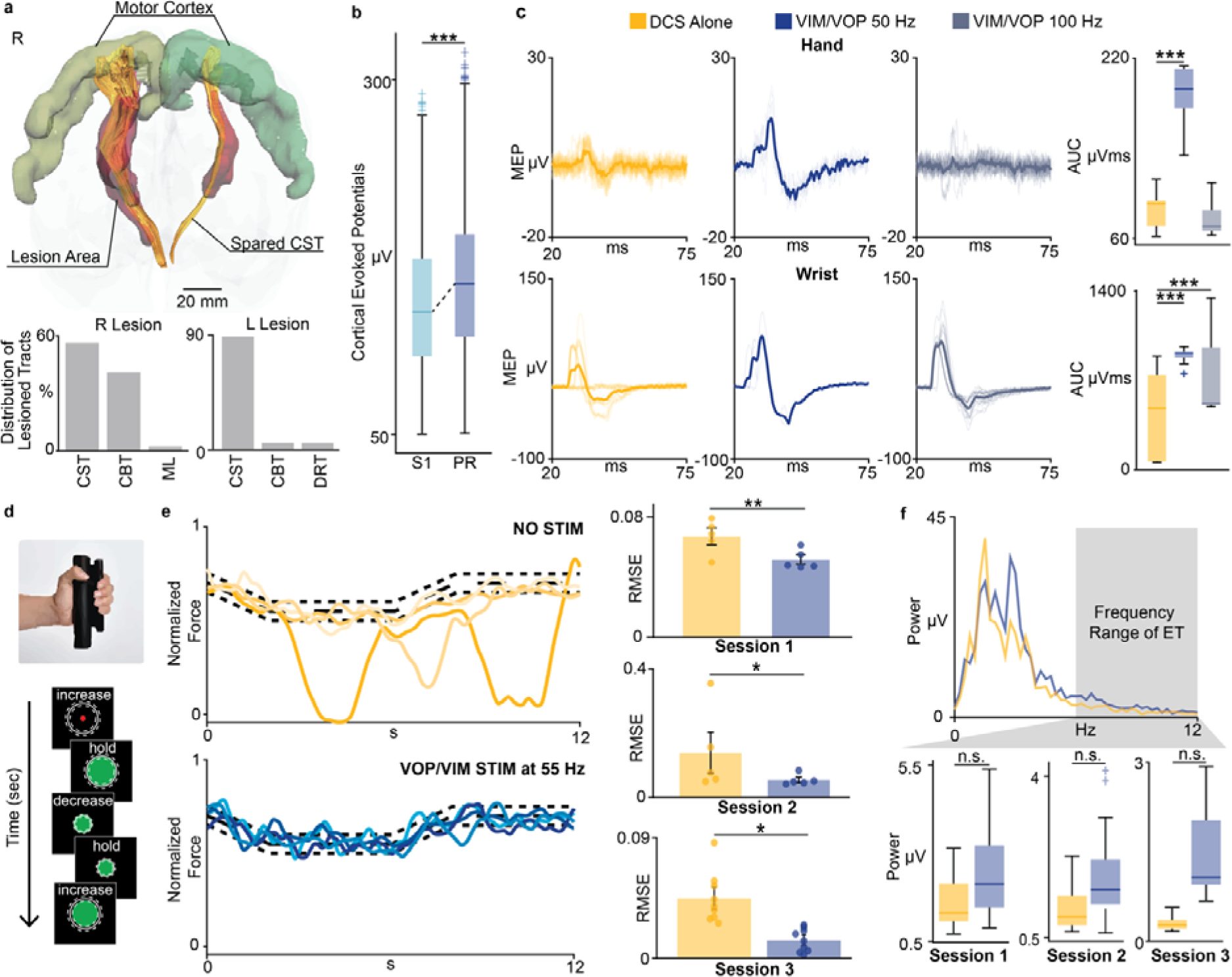
Stimulation of the motor thalamus improves voluntary motor control after lesions of the CST. **(a)** White-matter fibers damage estimation for CST01. *Top panel*: Differential tractography with regions of white matter tract lesion highlighted in red. *Bottom panels*: Proportion of lesioned tracts for left and right hemispheres (CST: corticospinal tract, CBT: corticobulbar tract, DRT: dentatorubrothalamic tract, ML: medial lemniscus). See **Extended Data Fig. 10** for lesion segmentation**. (b)** Box-plots of peak to peak amplitude of cortical EPs over S1 and PR contact from VIM/VOP stimulation at 10 Hz (n = 599). **(c)** *Left*: Example of MEPs with DCS alone and DCS paired with VIM/VOP stimulation at 50 and 100 Hz for fingers (APB) and hand (flexor) (n = 18). *Right*: box plots of MEPs AUC of ABP and flexor with DCS alone and DCS paired with VIM/VOP stimulation at 50 and 100 Hz. **(d)** Schema of the motor task performed by CST01. **(e)** *Left*: Example of force traces without (top) and with (bottom) VIM/VOP stimulation at 55 Hz. Yellow and blue intensity represent different repetitions. *Right*: bar plot of the root-mean-square error of the force without (yellow) and with (blue) VIM/VOP stimulation at 55 Hz (mean +/− standard deviation over trials). Statistical significance was assessed with two-tail bootstrapping with Bonferroni correction: p<0.05 (*), p<0.01 (**). **(f)** *Top*: average power spectrums from 1 to 12 Hz calculated over the hold periods of the task without (yellow) and whit (blue) stimulation at 55Hz. *Bottom*: boxplots of the average power spectrum over the clinical observed range for tremor (6-12Hz)^35^. No statistically significant difference was found between stimulation ON and OFF (one-tail bootstrapping with Bonferroni correction). For all boxplots, the whiskers extend to the maximum spread not considering outliers, central, top, and bottom lines represent median, 25^th^, and 75^th^ percentile, respectively.

### Stimulation of the motor thalamus improves voluntary force control in a person with CST lesion

Finally, to assess whether stimulation of the motor thalamus achieves functional improvements for chronic hemiparesis, we tested the effect of VIM/VOP stimulation in CST01 during a functional task using the chronically (> 5 months) implanted DBS system. Several previous works suggested that the CST is mostly responsible for dexterity and modulation of strength rather than for generation of bulk forces^34^. For this reason, we designed an isometric roadway test to measure voluntary force control. Specifically, CST01 was instructed to matched grip force to a time series of thresholds, gradually increasing (2 seconds), sustaining (4 seconds), and decreasing (2 seconds) the force between set percentages of maximum voluntary force levels that were established when stimulation was OFF (**Fig. 8d**). Tests were performed without and with bilateral VIM/VOP stimulation at 55 Hz. This experiment mimicked those performed in anesthetized monkeys (**Fig. 3c** and **Fig. 6c**). Importantly, the stimulation was set to 55Hz only during these tests. We repeated the testing over 3 different sessions to assess robustness of the immediate effects of the stimulation. On all three days, when the stimulation was turned ON at 55 Hz, CST01 was able to reduce the amount of grip force deviation from the requested force compared to when the stimulation was OFF. When the stimulation was ON performances were smoother and more accurate when compared to no stimulation (**Fig. 8e**, RMSE, Session 1: 0.05 (DBS ON) vs 0.07 (DBS OFF), Session 2: 0.05 (DBS ON) vs 0.14 (DBS OFF), Session 3: 0.013 (DBS ON) vs 0.045 (DBS OFF)). Importantly, when the stimulation was turned ON the patient did not report any side effects. Additionally, we controlled for the possibility that the improvement in motor performances was due to a reduction in tremor when the stimulation was ON. For this, we estimated the frequency power of the force trace and compared the average power from 6-12 Hz (frequency range typical of tremor)^35^ between stimulation ON and OFF. The average power was not different between stimulation ON and OFF proving that the tremor was consistent with and without stimulation at 55 Hz and demonstrating that the improved voluntary control was not caused by a reduction in the level of tremor (**Fig. 8f**).

While preliminary, these results suggest that stimulation of the motor thalamus at optimal stimulation frequencies (50-80 Hz) can improve volitional force control with the absence of noticeable side effects in patients with chronic lesions of the CST.

## Discussion

In this study we demonstrated with experiments in both monkeys and human subjects that stimulation of the motor thalamus facilitates the recruitment of cortico-spinal neurons within the motor cortex which in turn increases motor output in intact upper extremities and paretic limbs after lesions of the CST. Importantly, we elucidated the underlying mechanisms of action of this potentiation and consequently identified the optimal stimulation parameters to achieve maximal effect.

A few previous studies had already investigated the use of thalamic stimulation to treat paresis^36–40^. However, limited understanding of the underlying mechanisms of stimulation led to suboptimal choices of the implanted target and the stimulation protocols, which likely affected the size and consistency of the observed effects. Indeed, the widespread adoption of clinical DBS stimulation frequencies (> 100 Hz)^31^ prevented sustained MEP potentiation as those observed in our study with consequent reduced behavioral effects. Here, we built on these previous works, but we first investigated the mechanisms of action of motor thalamus stimulation to successfully tailor it to treat upper-limb motor deficits. Specifically, by combining intra-spinal and intra-cortical electrophysiology in monkeys and human subjects, we verified that stimulation of motor thalamus increased motor output by augmenting the recruitment of cortico-spinal motor neurons within the primary motor cortex via excitatory synaptic inputs from the targeted thalamic nuclei. Synaptic-mediated excitatory inputs are known to be less efficient at high stimulation frequency because of presynaptic inhibitory effects such as depletion of neurotransmitters and presynaptic inhibition^18,28,29^. For this reason, we explored subclinical DBS frequencies which led us to confirm that the 50-80 Hz range was optimal to sustainably increase motor output. Indeed, when trialing commonly used DBS stimulation frequencies (> 100 Hz)^31^ we found frequent *suppression* of motor output particularly of the fingers muscles, i.e., those innervated by the highest concentration of cortico-spinal tract axons^41^ and mostly affected by white matter lesions^42^. Importantly, for stimulation at high frequencies (i.e., > 100 Hz) we reduced the stimulation intensity to balance the total charge and verify that the suppression of the responses was indeed due to inhibitory mechanisms and not a confound from a difference in charge delivered.

This mechanisms-driven selection of the target and parameters of stimulation resulted in a sustained MEP potentiation with an average of 500% increase over all muscles and subjects, and which led to enhanced grip force control. Indeed, stimulation of the motor thalamus availed a patient with a severe, chronic lesion of the CST to volitionally modulate grip force, demonstrating that continuous stimulation of the motor thalamus improved fine motor control. Previous studies using non-invasive brain stimulation such as TMS or transcranial direct-current stimulation (tDCs) already proposed that enhanced cortical excitability could improve motor functions after CST lesions^43–51^. However, non-invasive stimulation induced significantly smaller effect sizes in MEPs potentiation compared to the average 2-fold increases that we reported here. We speculate that these large effect sizes are necessary to observe the immediate therapeutic effects in motor control that we measured in our participant when VIM/VOP-DBS was ON. Importantly, we believe that these effect sizes are caused by the high degree of selectivity of our approach that enables powerful, but selective excitatory drive to motor cortex ^52,53^. Importantly, we obtained large behavioral effects by simply activating the stimulation. However, previous studies in brain stimulation^43–51^ suggest that combining stimulation with physical training may lead to the emergence of long-term therapeutic changes that could further increase effect sizes highlighting a path to combine DBS of the VIM/VOP with protocolled upper limb rehabilitation.

These results offer promising albeit preliminary support for the clinical use of DBS of the VIM/VOP as a therapy to improve motor deficits in people with lesions of the CST. Importantly, in the tested patients we did not find side effects on speech, gait difficulties, and ataxia, which are often clinically observed with VIM/VOP stimulation. Importantly, VIM/VOP stimulation side-effects are usually reported with clinical standard frequencies (>100 Hz). We, therefore, believe that the use of lower frequencies (i.e., 50-80Hz) might limit these secondary effects enabling a safe and effective use of DBS for motor improvements. A potential limiting factor for widespread clinical use of DBS in the context of motor deficits induced by lesion of the CST is surgical safety. In fact, adverse events of DBS implantation are considered low (<0.5% of patients)^54^ and are balanced by the potential benefit gain. Moreover, surgical risk in patients with a history of cerebral lesions can be minimized by careful pre-operative management of anticoagulants and by delaying the implantation for at least three months following the brain insult^55,56^. This was recently demonstrated in a study applying DBS to the cerebellar dentate nucleus in n = 12 individuals^57^ and in a large clinical trial (n=108) implanting vagus nerve stimulators to treat post-stroke motor deficits^58^. Therefore, the surgical intervention required for DBS implantation does not represent a significant barrier to its clinical translation to treat patients with CST lesions. Additionally, the VIM/VOP target is commonly used as a target for DBS therapy in clinical practice for the treatment of Essential Tremor, and the devices used in this study are commercially available, FDA approved, and currently implanted in more than 12,000 patients per year for the treatment of different movement and psychiatric disorders^52^. DBS has changed the treatment options for millions of patients with motor disorders such as Parkinson’s Disease and Essential Tremor making the perspective of an application in people with white matter lesions highly promising and appealing for clinicians^36^. Yet, our approach proposes a new principle of DBS, i.e., the use of DBS not to inhibit but to enhance motor functions. Additionally, our results demonstrate the general principle that stimulation of subcortical regions projecting to the motor cortex could increase cortical excitability and consequently potentiate motor output. While DBS could be a viable approach for this, novel non-invasive techniques such as focused ultrasound or transcranial electrical temporal interference could overcome the surgical limitations imposed by DBS potentially serving a larger patient population^59–61^. Additionally, other subcortical targets with excitatory projections to the motor cortex, such as the centromedian thalamic nucleus, could have effects similar to those reported here and would require future investigations.

The most important limitation of this study is that the behavioral tests in humans were performed in only one patient with CST lesion. However, the main goal of this work was to first identify the mechanisms of action of thalamic stimulation to optimize stimulation targets and parameters and quantify the immediate assistive effects. We extensively performed these investigations in a large number of monkeys and human subjects demonstrating the consistency of our findings across species, including one patient suffering from a chronic (> 6 months) CST lesion. This consistency across monkeys and human subjects importantly emphasizes that the observed MEP potentiation is present regardless of etiology and is not due to a change in cortical excitability secondary to the tremor^62–64^ further supporting tests in a larger cohort of patients with CST lesions.

In conclusion, here we optimized motor thalamus stimulation to enhance motor outputs setting the stage towards a novel effective therapy for motor related deficits after white matter lesions. With a series of experiments progressing from monkeys to human subjects, we showed that targeted stimulation of thalamocortical excitatory pathways to cortico-spinal motoneuron could increase their excitability restoring voluntary motor control. Interestingly, these mechanisms resemble those previously proposed for SCS. Indeed, we recently showed that SCS of the cervical spinal cord improved upper-limb strength and volitional control by directly activating sensory afferents in the dorsal roots, which form mono- and polysynaptic excitatory connections to spinal motoneurons, and thus increasing the responsiveness of the spinal motoneurons to the remaining CST inputs^65,66^. Paralleling these two results, we could speculate a general principle that might apply to other areas of the central nervous system and could guide other neuromodulation applications: direct stimulation of excitatory pathways with strong connectivity to a target area of interest (e.g. motor cortex) improves neural function (e.g. motor output) without disrupting ongoing neural activity. Future studies in patients with cerebral white matter lesions are now necessary to further demonstrate safety and efficacy of our approach for the recovery of fine motor control.

## Methods

### Animals

All procedures were approved by the University of Pittsburgh Institutional Animal Care and Use Committee (protocol number IS0017081). We utilized 4 adult Macaca Fascicularis (3 male: MK-SC 4y.o. 5.4kg, MK-SZ 4y.o. 5.9kg, MK-JC 6y.o. 7.5kg, 1 Female: MK-OP 6y.o. 6kg) and 1 adult male Macaca Mulatta (MK-HS 7y.o. 12kg). The animals were housed in the Division of laboratory Animal Resources at the University of Pittsburgh. When possible, the animals were pair housed with another animal prior to any surgical procedures. The animals were not water or food restricted and were given daily enrichments (novel food, toys, puzzles). Detailed information on which animals were involved in specific experimental procedures are reported in **Supplementary Table 1**.

### Animal surgical procedure

For each animal, we performed one survival surgical procedure and one terminal surgical procedure. During the survival surgical procedure, we implanted five fiducial titanium screws (1.5mm x 4mm, KLS Martin) in the skull at non-coplanar depths. We then performed a computed tomography (CT) scan (250μm isotropic, Epica Vimago GT30) of the skull with the implanted screws. The procedure was performed using standard aseptic techniques under full anesthesia induced with ketamine (10 mg/km, i.m.) and maintained under isoflurane (1-3%, 2 L/min, inhalant). Over the next 7 days, we injected anti-inflammatory drugs once per day (Rimadyl 4mg/kg, Dexamethasone 0.4 mg/kg).

During the terminal surgery the following procedures were performed: 1) peripheral nerve and muscle implantation, 2) robotic deep probe implantation, 3) intra-cortical electrodes arrays implantation, 4) spinal probe implantation, and 5) subcortical lesioning. These procedures were performed under full anesthesia induced with ketamine (10 mg/kg, i.m.) and maintained under continuous intravenous infusion of propofol (1.8-5.4 ml/kg/h) and fentanyl (0.2-1.7 ml/kg/h). Anesthesia affects neural responses and local excitability. However, propofol is known to minimize these effects and it is often used in electrophysiology^67^. Certified neurosurgeons performed these surgical procedures (Drs. Jorge Gonzalez-Martinez, Peter Gerszten, Daryl Fields, Vahagn Karapetyan, UPMC, Pittsburgh, USA). At the end of the experiments, the animals were euthanized with a single injection of pentobarbital (86 mg/kg) and perfused with 4% paraformaldehyde (1 L/kg) for further tissue imaging.

#### Peripheral nerve and muscle implantation

We dissected from the lateral epicondyle of the arm and implanted a cuff electrode (FNC-2000-V-R-A-30 bipolar nerve-cuff Micro-Leads Neuro, Ann Arbor, MI, USA) around the deep branch of the radial nerve (motor branch). We electrically stimulated two branches of the radial nerve and assessed the EMG response to verify the motor branch from the cutaneous branch. We then inserted EMG needle electrodes (disposable single 13mm Subdermal needle electrode, Rhythmlink) into the extensor carpi radialis (ECR), extensor digitorum communis (EDC), flexor digiti minimi (FDM), flexor carpi radialis (FCR), flexor digitorum communis (FDC), abductor pollicis brevis (APB), and biceps (hand and arm muscles). Additionally, in n=3 animals (MK-OP, MK-HS, MK-JC), we implanted EMG electrode needles in the Masseter (Mas), Orbicularis Oris (O. oris), and Buccinator (Buc) (face muscles).

#### Robotic deep probes implantation

We implanted each depth electrode using the ROSA One(R) Robot Assistance Platform (ROSA robot, **Extended Data Fig. 1**) to allow for highly accurate implantations of the ventral laterolateral (VLL) thalamic nucleus and the hand area of the internal capsule (IC)^68^. Prior to the terminal surgery, we obtained preoperative T1-weighted magnetic resonance imaging (MRI) scan (400μm isotropic, TR/TE: 6000/3.7 ms, 7T Siemens whole body human system) and CT imaging to estimate precise targets and trajectories for each electrode. Both imaging procedures were performed with the animals secured in a customized plastic stereotactic frame to facilitate co-registration. We then co-registered MRI and CT images using ROSA One Brain Application and selected three targets: the hand area of the cortico-spinal tract (CST) within the IC at the AC-PC level (stimulating electrode), a target 2 cm ventral to create a thermal-ablation lesion (lesioning electrode), and the VLL nucleus of the motor thalamus. We planned the trajectories of the probe in the ROSA software, avoiding vasculature and the ventricles. The entrance of the probes was positioned 2 cm in front of the central sulcus to keep the motor and somatosensory cortices intact for the implantation of the intra-cortical electrode arrays. This entrance required an implantation trajectory with a different angle for each probe. The ROSA robot was, therefore, critical to avoid a crossing of the three probes. We then positioned the monkeys prone in a stereotaxic head frame (Kopf, Model 1530, Tujunga, CA, USA) and registered the ROSA robot to the monkey using the implanted fiducial screws (see **Extended Data Fig. 1** for precision of the co-registration). The robot guided the drilling of penetration holes and the precise implant of the fixation bolts. For the two probes within the internal capsule, we inserted and fixed a radiofrequency cannula (S-100 5 mm ActiveTip Straight cannula 22G, Abbott) and a radiofrequency electrode (RF-SE-10 Reusable Stainless-Steel Electrode, Abbott) at the correct depth into the brain. Correct positions of the probes within the IC were estimated by recording evoked electromyography (EMG) potentials from stimulation of the IC at 2 Hz at amplitudes between 800uA and 2mA that should produce mono-synaptic activation of the cervical motoneurons. For the stimulation of the VLL, we inserted a 16 channel Dixi electrode (DIXI Microdeep® SEEG Electrodes) into its fixation bolt. Correct position within the VLL was confirmed by recording evoked potentials in the cortex during electrical stimulation (1 Hz) at amplitudes between 1 and 4.8mA that elicited clear evoked potentials in the motor cortex, but not in the somatosensory cortex. Detailed information on the amplitudes and pulse durations used for stimulation of the IC and the VLL are reported in **Supplementary Table 2.**

#### Intra-cortical electrodes arrays implantation

We performed a 20 mm diameter craniotomy over the central sulcus and removed the dura to expose primary motor and primary somatosensory cortices. Functional motor areas of the arm were identified through anatomical landmarks and intra-surgical micro-stimulation. The position of the primary somatosensory area (S1) was then determined in relation to the hand representation of the primary motor cortex (M1). We then implanted intracortical arrays in M1 (48 channels) and S1 (48 channels) for a total of 96 channels (400μm pitch, electrode length 1.0mm Blackrock Microsystems, Salt Lake City, UT, USA). In MK-JC, we implanted a 64-channel array into M1 and a 48-channel array into S1 (total 112 channels, 400μm pitch, electrode length 1.0mm Blackrock Microsystems, Salt Lake City, UT, USA). The arrays’ implantation was achieved using a pneumatic compressor system (Impactor System, Blackrock Microsystems).

#### Spinal probe implantation

After the completion of the cranial surgery, we performed a laminectomy from C3 to T1 vertebrae and exposed the cervical spinal cord. We then implanted a 32-channel linear spinal probe (A1×32-15mm-100-177-CM32 Linear Probe with 32 pin Omnetics Connector, NeuroNexus, Ann Arbor, MI, USA) in the gray matter at the C6-C7 spinal segment to record spinal local field potentials and multi-unit spikes. To implant the probe, we opened the dura mater and placed a small hole in the pia using a surgical needle through which penetration of the probe with micromanipulators was possible. We implanted the probe using Kopf micromanipulators (Kopf, Model 1760, Tujuna, CA, USA).

#### Subcortical lesioning

We utilized a radiofrequency generator (NeuroTherm NT 1100) to create the lesions^32^. Time and temperature parameters used for the lesion for each animal are summarized in **Supplementary Table 3.** At the end of the experiments before perfusion, we also created a small lesion (60E for 5 seconds) in VLL through the Dixi electrode to visualize the implant location post-mortem.

### Data acquisition and electrophysiology

Stimulation of the IC, VLL, and radial nerve was provided using an AM stimulator (model 2100 A-M Systems, Sequim, WA, USA). Stimulation was delivered as either single pulses or bursts of cathodal, charge balanced, symmetric square pulses. The stimulation intensity for the IC and the nerve was set for each animal at the motor threshold (i.e., when movements became visible). Detailed information on the amplitudes and pulse widths used for stimulation of the IC, the VLL, and the radial nerve are reported in **Supplementary Table 2.**

All electrophysiological and neural data was amplified, digitally processed, and recorded using the Ripple Neuro Grapevine and Trellis software at a sampling frequency of 30000 Hz. Neural events were determined for each channel of the Utah arrays and linear probes by applying a broadband filter between 300 Hz and 3 kHz and setting a voltage threshold of 3 root-mean-square. LFPs were filtered between 0 and 500 Hz and then downsampled to a sample frequency of 1000 Hz.

### Grasp force data acquisition

To produce a grasp motion from an anesthetized monkey (MK-JC), we stimulated the IC every two seconds with a 1 second pulse train at 47 Hz and 1 mA. We collected the grasp force data using a 6-axis low-profile force and torque (F/T) sensor (Mini40, ATI Industrial Automation, North Carolina). The sensor was powered using a multi-axis (F/T) transducer system (ATI DAQ F/T) that also calibrated the force data. The sensor was able to measure three degrees of force (+/-810 N for Fxy, +/-2400 N Fz) and torque (+/− 19Nm Txy, +/− 20Nm Tz) simultaneously. A small rod was mounted to the xy-plane of the sensor to be a grip handle for the animal and the system was secure under the animal’s left arm. The force and torque data were digitized and recorded using the Ripple Neuro Grapevine and Trellis software at a sampling frequency of 30000 Hz.

### Animal post-mortem High Definition Fiber Tracking (HDFT)

After perfusion, the brain was dissected for post-mortem imaging and soaked with 0.2% gadolinium (1 mmol/mL, Gadavist, Bayer) in 1x PBS for 2 weeks and then with 0.2% gadolinium in pure water for 2 days^69^. The brain was then placed in a 3D customized 3D-printed holder filled with Fomblin for tissue magnetic susceptibility matching and to prevent specimen dehydration during scanning. We then acquired T2-weighted MRI (125 μm isotropic resolution, TR/TE: 1500/60.57 ms) and diffusion-weighted MRI (0.5 mm isotropic resolution, b-values=1800, 3400, and 5200 s/mm^2^, with 30, 40, and 90 diffusion directions, respectively, and 6 A_0_ images). Image data were acquired using a 9.4T/31cm horizontal-bore Bruker AV3 HD animal scanner equipped with a high-performance 12-cm gradient set, capable of 660 mT/m maximum gradient strength, and a 72mm quadrature birdcage RF coil. Diffusion tensor estimation and tractography were performed using DSI studio (http://dsi-studio.labsolver.org). The accuracy of b-table orientation was examined by comparing fiber orientations with those of a population-averaged template^70^. The restricted diffusion was quantified using restricted diffusion imaging^71^. The diffusion data were reconstructed using generalized q-sampling imaging^72^ with a diffusion sampling length ratio of 0.6.

For fiber tracking to characterize the thalamocortical projections, we used a tracking threshold of 0, angular threshold of 0, and a step size of 1 mm. Tracks with lengths shorter than 20 mm or longer than 200 mm were discarded^73^. This ensured that all possible tracts were captured while reducing the amount of tract fragments similar to Ghulam-Jelani et al., 2021^74^. A total of 10,000 tracks were placed. Topology informed pruning^75^ was applied to the tractography with 2 interactions to remove false connections. We selected each of three regions of the monkey motor thalamus (the ventral anterolateral region (VAL), ventral laterolateral region (VLL), and ventral posterolateral region (VPL)) as a seed to create tracks that projected to the cortical areas of interest: the primary somatosensory cortex (S1), the primary motor cortex (M1), the dorsal premotor cortex (PMd), and the supplementary motor area (SMA). Cortical and subcortical regions were mapped into each animal MRI using the built-in primate CVIM atlas^76^ and confirmed by certified neurosurgeons (Dr. Jorge Gonzalez-Martinez). For each nucleus (VAL, VLL, and VPL), we calculated the volume of the projections to each cortical region of interest (S1, M1, PMd, and SMA). We then normalized each projection volume by the sum volume of all projections from that thalamic nucleus. We repeated this analysis for all thalamic nuclei individually. This allowed us to estimate the proportion of fibers that projects to a cortical area from a single thalamic nucleus and to compare the proportion of projections to each cortical area between the different thalamic nuclei. To test the robustness of our results, we repeated the analysis using different tractography parameters (i.e., tracks discarded if lengths shorter than 25mm or longer than 250mm and a total of 12,000 tracks) and confirmed that the results were the same (data not shown).

To analyze the selectivity of projections to only M1 from each of the motor thalamus nuclei, we computed a receiver operating characteristic (ROC) curve comparing the true positive rate and false positive rate of projections to M1 compared to those not to M1. We counted both rates while thresholding at each unique normalized volume. We then computed the area under the ROC curve for the VPL, VLL, and VAL.

To quantify the projections from the area of stimulation of the depth electrode (i.e., electrode within or in close proximity to the VLL), we manually drew a region, based on the size of the thermal-ablation lesion. The thermal-ablation was created at the end of the experiments with the objective to visualize the location of the depth electrode post-mortem. We then mirrored this region to the intact contralateral side (due to disruption of thalamocortical fibers by the lesion) and selected it as the seed. S1, M1, PMd, and SMA were selected as regions of interest. We used the same tracking parameters as above. We then calculated the volume of each tract normalized by the total volume of tracts from the area of stimulation to all cortical areas.

To quantify the damaged cortico-spinal tract (CST), we used a tracking threshold of 0, angular threshold of 0, and a step size of 0 mm. Tracks with lengths shorter than 30 mm or longer than 500 mm were discarded^73^. This ensured that all possible tracts were captured while reducing the amount of tract fragments similar to Zhang et al., 2021^77^. A total of 10,000 tracks were placed. Topology informed pruning was applied to the tractography with 2 interactions to remove false connections. To reconstruct the CST, we manually drew regions of interest in the left and right cerebral peduncles (confirmed by neurosurgeon, Dr. Jorge Gonzalez-Martinez). We considered the respective left and right sensorimotor cortical areas (M1, S1, PMd, ventral pre-motor cortex – PMv, and SMA) as seed regions. We then calculated the volume of each CST and normalized it by the volume of the intact CST.

### Analysis of cortical activity

The broadband cortical data was first bandpass filtered between 10 and 5000 Hz with a 3rd order Butterworth filter and 1ms blanking was applied over stimulation artifacts. We then extracted a 25 ms window (5ms before stimulation, 20ms after stimulation) to capture the entirety of the cortical evoked potential. We then calculated the peak to peak amplitude and averaged it across all stimulation trials. We quantified multi-unit activity offline by calculating the average spike count across all trials for each channel. Cortical spikes were detected with a threshold of 3-3.5 (MK-SC: 3, MK-SZ: 3, MK-OP: 3.5, MK-HS: 3, MK-JC: 3) standard deviations above baseline for each channel of the 2 Utah arrays. The average spike counts were baseline corrected and were calculated using a bin size of 2 ms. We blanked 13 ms after stimulation to remove stimulation artifacts.

To analyze antidromic potentials to IC stimulation, we recorded local field potentials in the M1 Utah array when stimulating the IC at 2 Hz. We then sent a 100 ms 100 Hz burst of stimulation to the VLL preceding a single pulse of IC stimulation at various delays (2, 5, 10, and 50 ms). We applied the same filtering to the cortical data as above. We extracted a 45 ms window (5 ms before IC stimulation, 40 ms after IC stimulation) to capture the entire antidromic potential. We then calculated the peak to peak amplitude of the antidromic potential and averaged it across all stimulation trials.

To calculate the frequency dependent cortical responses to VLL, we took the bandpass filtered cortical evoked potentials over M1 and extracted 12 ms windows (2 ms before stimulation, 10 ms after stimulation) for each stimulation pulse at a variety of VLL frequencies (VLL stim 10, 50, 80, and 100 Hz). We then calculated the peak to peak amplitude of the evoked potential over the time range of 1.5 to 10 ms after the simulations and averaged it across all stimulation trials.

### Analysis of muscle activity and kinematics

Electromyographic activity was bandpass filtered between 30 and 800 Hz with a 3rd order Butterworth filter. We then computed the stimulation triggered averages (window from 5 ms to 25 ms after IC stimulation) of motor evoked potentials and calculated the area under the curve (AUC) for each pulse of IC stimulation and each muscle.

Kinematics were recorded by a GoPro® Camera and analyzed through Deeplabcut^78^, a neural network for 2D and 3D markerless pose estimation based on transfer learning. A subset of video frames (40-80 frames) per condition were selected by Deeplabcut to capture diversity of the movement. Next, we manually labeled the thumb, index finger, wrist, and elbow in each of those frames. With the labeled frames, we used Deeplabcut to create a training network by merging all extracted labeled frames. We then used the trained network to analyze all the videos and extract the kinematic traces for each of the labeled body points. We calculated the peak to peak amplitude in pixels of each joint for each pulse of IC stimulation.

### Analysis of frequency dependent modulation of muscle responses

For each animal, muscle, and IC paired VLL stimulation (10, 50, 80, 100, and 200 Hz) protocol we concatenated all MEP responses from each IC stimulation pulse (window from 0 ms to 25 ms after IC stimulation). Importantly, stimulation amplitude at 200 Hz was reduced to balance the total charge (see **Supplementary Table 2** for stimulation parameters). We then used a 5th-order polynomial regression fit model to classify these MEP traces according to four criteria^28^: 1) “no potentiation” - the responses are consistent with IC alone stimulation, 2) “sustained potentiation” - the responses are increased as compared to IC alone, 3) “attenuation” - the responses decrease through time, 4) “suppression” - the responses are suppressed through time. The categorization was validated by visual inspection from two independent evaluators. For this, the evaluators were blinded to the stimulation conditions and to the muscles name when inspecting and categorizing the MEP traces. For each unique stimulation protocol, we calculated its probability distribution over all muscles and animals.

### Analysis of spinal activity

We applied the same filtering and extraction process to the spinal evoked potentials as we did to the cortical evoked potentials (filter 10-5000 Hz 3rd order Butterworth and 25 ms windows). To identify the antidromic response from the VLL stimulation, we calculated the peak to peak amplitude and response onset within the first 5 ms following VLL stimulation across all channels of the intraspinal probe. Using the intraspinal probe map we were able to distinguish the ventral, intermediate, and dorsal areas on the probe. In order to compare the responses across animals, we normalized the peak to peak amplitude by the maximum value for each animal independently.

When assessing the frequency dependent effects of VLL stimulation paired with IC stimulation on the spinal responses, we blanked the IC stimulation artifact between 250 us before stimulation and 500 us after the stimulation pulse ended. We then calculated the area under the curve (AUC) of the spinal evoked potentials for each VLL stimulation protocol (10, 50, 80, and 100Hz). The AUC were calculated from 5 to 10 ms after IC stimulation to exclude any stimulation artifacts and secondary post-synaptic responses. The AUC were compared against the AUC of the spinal responses from IC stimulation alone for potentiation or attenuation.

### Human participants

We performed electrophysiological experiments on n=4 human subjects (2 males and 2 females) of age 69.5 ± 8.54 (mean±std) who were undergoing DBS implantation of the VIM/VOP nucleus (ventralis intermediate nucleus/ventralis oralis posterior), which corresponds to the VLL nucleus in monkeys, to treat medically-intractable asymmetric Essential Tremor (ET) symptoms. We tested the least symptomatic arm to better approximate normal function. Additionally, we tested a traumatic brain injury (CST01) patient (male in his 40’s) who had suffered subarachnoid hemorrhaging from a motor vehicle accident. This hemorrhage led to diffuse axonal injury involving the CST and bilateral edema of the cerebral peduncles and pons (**Fig. 8a** and **Extended Data Fig. 10**, more pronounced in the left than the right hemisphere). Consequently, CST01 suffered from hemiparesis of the right side and left upper extremity muscle weakness and tremor with decreased overall strength, balance, coordination, and endurance. He required maximal assistance for ambulating (taking steps, shifting weight) and daily tasks requiring dexterous control (splint/bandage changes, eating, showering, oral hygiene). He was able to sit without support, stand from a sitting position with moderate assistance, and follow oral commands. The patient was recommended for bilateral DBS electrode implant to treat the post-traumatic tremor. All intra-operative procedures were approved by the University of Pittsburgh Institutional Review Board (STUDY21040121). The behavioral experiments were approved by the University of Pittsburgh Institutional Review Board (STUDY21100020).

### Human intraoperative data acquisition and electrophysiology

Pairs of needle electrodes (Rhythmlink Columbia, SC) were placed subcutaneously over the deltoid, biceps brachii, triceps brachii, flexors (flexor carpi ulnaris, FLEX), extensor (extensor carpi radialis longus, EXT), and abductor pollicis brevis (APB) muscles to record MEPs. Prior to DBS insertion, a subdural strip electrode (6 contact platinum subdural electrode, AD-TECH Medical Instrument Corporation, Oak Creek, WI) was implanted over the cortical surface with verification of positioning provided using median nerve somatosensory evoked potential (SSEP) phase reversal (PR) mapping to locate the hand representation of S1 cortex and the approximate location of the central sulcus^33^. The electrode position was optimized using results from direct cortical stimulation (DCS) MEPs to contralateral upper extremity muscles. We located the contact over the precentral gyrus that generated the largest amplitude MEP in the hand muscle (APB). DCS of the hand representation of M1 was provided using trains of 5 stimulation pulses (0.5 ms) at 400 Hz every two seconds at stimulus intensities up to 15mA using an intraoperative neurophysiological monitoring system (XLTEK Protektor, Natus Medical, Ontario, Canada). Detailed information on the stimulation parameters used for DCS for each subject are reported in **Supplementary Table 4.** After implantation of the subdural strip electrode, the surgery continued following standard clinical procedures and DBS electrodes were implanted in the VIM/VOP nucleus bilaterally using subcortical mapping techniques. The electrophysiology experiments were performed following completion of the clinical procedure, with the subject maintained under sedation with propofol. Specifically, DCS MEPs from stimulation of the optimal electrode contact over the hand representation of the primary motor cortex were recorded without and with continuous stimulation of the VIM/VOP nucleus (DBS contacts −1 +8) at 50, 80, and 100Hz with pulses of 100us and intensity of 3mA. MEPs were collected for 100 ms trials following each DCS stimulation burst and recorded using the XLTEK system at a sampling frequency of 6000 Hz. Stimulation of the VIM/VOP was delivered via the Boston Scientific clinician programmer that connects via Bluetooth to an external trial stimulator. The same DBS electrode was used to generate cortical evoked potentials in the subdural strip electrode that were recorded using the Ripple Neuro Grapevine and Trellis software at a sampling frequency of 30000 Hz.

### Behavioral experimental recordings and analysis

To measure the level of voluntary force control, the participant was asked to grasp a force dynamometer (Pinch/Grip Digital Myometer, MIE Medical Research, Yorkshire, UK), which recorded the force in newtons at a sampling rate of 30000 Hz. The participant was tasked to modulate their force between two limits (55-70% of maximum voluntary contraction measured with stimulation OFF) while receiving visual feedback of the target force and the applied force on the screen. The visual feedback of the target force curve consisted of a white circle that changed in size in relation to the expected target force. The patient had to keep the force within +/-5 % of the expected target force (visually displayed as dashed boundaries around the white circle). A second circle represented the force exerted by the patient and it would be green if the force was within the +/-5% or red if outside this boundary. The target force curve started at the high force level (70% of MVC) followed by a linear decrease for 2 seconds, followed by a hold phase at the low force level (55% of MVC) for 4 seconds, followed by a linear increase for 2 seconds, and finally a hold phase at the high force level for 4 seconds. This cycle was repeated 4 times. We calculated the root-mean-squared error between the applied force and the expected force at each time point.

To calculate the frequency power of the force trace, we filtered with a band-pass 2nd order Butterworth filter (cut-off frequencies of 2-5000 Hz) and performed a fast fourier transform of the hold portion of the force task. We then compared the average power from 6-12 Hz (frequency range typical of tremor)^35^ during VIM/VOP stimulation on and off.

### Analysis of intraoperative cortical and muscle activity

Electrocorticographic data were filtered with a band-pass 2nd order Butterworth filter (cut-off frequencies 10-500 Hz) and a 60 Hz notch 2nd order Butterworth filter. The stimulation artifact was blanked. We extracted epochs of 30 ms (10 ms before and 20 ms after the stimulus onset) and computed stimulation triggered averages. We calculated the peak to peak amplitude of each cortical evoked potential as the difference between the maximum and minimum voltage value in the interval 3-15 ms following the stimulus onset.

To quantify DCS MEPs recorded from triggered electromyographic activity, we computed the stimulation triggered averages (EMG from 10 ms to 75 ms after stimulation) and calculated the AUC for each muscle.

### Human Volume of Activation, HDFT, and Lesion Segmentation

#### Volume of Activation

After DBS implantation of the VIM/VOP DBS electrode, patients underwent post-implant CT imaging (1.25 mm isotropic, GE Medical Systems) to reconstruct the DBS leads. With the reconstructed DBS leads, we modeled the volume of tissue activation (VTA) in each patient using the DBS analysis module inside BrainLab Quentry®. Specifically, we visualized a 3D reconstruction of VIM, DBS electrode, and the simulated VTAs from the anode and cathode of the DBS leads. We then aggregated the VTA from the DBS patients (excluding CST01) and overlaid them on axial and coronal T2-weighted MRI images (1.6 mm isotropic, TR/TE = 9690/91 ms, 3T Siemens Prisma Fit).

#### HDFT

The diffusion images were acquired on a SIEMENS Prisma Fit scanner using a diffusion sequence (2mm isotropic resolution, TE/TR= 99.2 ms/2490 ms, 257 diffusion sampling with maximum b-value 4010 s/mm²). The accuracy of b-table orientation was examined by comparing fiber orientations with those of a population-averaged template^70^. The tensor metrics were calculated using DWI with b-value lower than 1750 s/mm².

To quantify the projections from the area of stimulation within the VIM/VOP DBS electrode, we manually drew a region at the stimulation contact and considered it as the seed region. S1, M1, PMd and SMA were the regions of interest. For fiber tracking, we used a tracking threshold of 0, angular threshold of 50, and a step size of 1 mm. Tracks with lengths shorter than 30 mm or longer than 500 mm were discarded. A total of 10,000 tracks were placed. Topology informed pruning was applied to the tractography with 2 interactions to remove false connections. We then calculated the volume of the tracts between the seed region and each region of interest normalized by the total volume of tracts from the area of stimulation to all cortical areas.

To quantify the white-matter fibers damage in CST01, we used an advance diffusion MRI technique commonly used in patients with bilateral white-matter damage that allows for quantification of a decrease in anisotropic diffusion by applying a “tracking-the-difference” paradigm when compared to a normal population template: differential tractography^79^. Differential tractography was performed by comparing the HDFT of CST01 against an age-and-sex-matched template constructed from a group of healthy controls (approximately 40 years old and male) of healthy quantitative anisotropy (QA). To capture neural injury, we highlighted tracks within CST01 that displayed a 20% decrease in QA when compared to the template of healthy controls. We then quantified the composition of lesioned tracts based on a population-averaged atlas of human white matter tracts^70^ for the main fiber tracts: corticospinal tract, corticobulbar tractdentatorubrothalamic tract and medial lemniscus.

#### Lesion Segmentation

In CST01, we highlighted areas of lesion. Lesion segmentation was performed by board-certified neurosurgeon (Dr. Jorge Gonzalez-Martinez) from axial T2-flair images acquired by a 3T Siemens Prisma Fit Scanner (1.6 mm isotropic, TR/TE = 9690/91 ms).

### Statistical Procedures

All statistical comparisons of means presented in this manuscript were performed using the bootstrap method, a non-parametric approach which makes no distributional assumptions on the observed data. Instead, bootstrapping uses resampling to construct empirical confidence intervals for quantities of interest. For each comparison, we construct bootstrap samples by drawing a sample with replacement from observed measurements, while preserving the number of measurements in each condition. We construct 10,000 bootstrap samples and, for each, calculate the difference in means of the resampled data. We employed two tailed bootstrapping with alphas of 0.05 (95% confidence interval), 0.01 (99% confidence interval), or 0.001 (99.9% confidence interval). The null hypothesis of no difference in the mean was rejected if 0 was not included in the confidence interval of the corresponding alpha value. If more than one comparison was being performed at once, we used a Bonferroni correction by dividing the alpha value by the number of pairwise comparisons being performed.

### Reporting Summary

Further information on research design is available in the Nature Research Reporting Summary linked to this article.

## Data availability

The main data supporting the results in this study are available within the paper and its Supplementary Information. All data generated in this study and software will be uploaded in a public repository upon acceptance of the manuscript. Raw data will be available upon reasonable request to the corresponding author.

## Acknowledgements

We thank Isabella Bushko for the design of fig. elements, Drs. Peter Strick and Aaron Batista for support with monkey housing, Dr. Frank Yet for providing support with the differential tractography analysis, Dr. Amr Mahrous for providing support during the monkey surgeries, and Christopher Pappas for helping with the VIM/VOP stimulation during the human surgeries. We wish to thank Zimmer Biomet for lending the ROSA robot to use in our surgeries (we declare no conflict of interest). We would like to thank Merek Gourley for providing post-implant reconstructions of the DBS patients.

## Author contributions

EP and JGM conceived the study. EP, JGM, and MC secured funding. JH, EG, DC, MC, JGM, and EP designed the experiments. JH, EG, JB, LL, MC, and EP designed and implemented hardware and software for the monkey experiments. JH and LL performed the image processing for all the monkeys’ experiments. TKH collected the monkeys’ imaging data. JH, LL, and JBM processed the DTI data for both the human and the monkeys. JH and SK processed the deep lab cut data for the kinematic analysis of the monkeys. JH, LL, JGM and EP designed the neurosurgical approach for the brain implants and JGM performed the surgery. JH, LL, JB, PG and MC designed the neurosurgical approach for the spinal implants and PG performed the surgery. VK and DF assisted with all monkeys’ surgeries. TC and JGM implemented patient recruitment, eligibility and monitoring and coordinated study management. JH, EG, AD, and EP collected all human and monkey data with the assistance of JB, LL, and MC for the monkeys’ experiments. DC performed the subcortical mapping of VIM/VOP for the human DBS implantations. GMA and DC collected the electrophysiology human data. JH, EG and AD analyzed the data. EP, JGM, MC and DC helped with data interpretation. EP, JGM, JH, EG and AD wrote the paper and all authors contributed to its editing.

---

**Extended Data Fig. 1:**
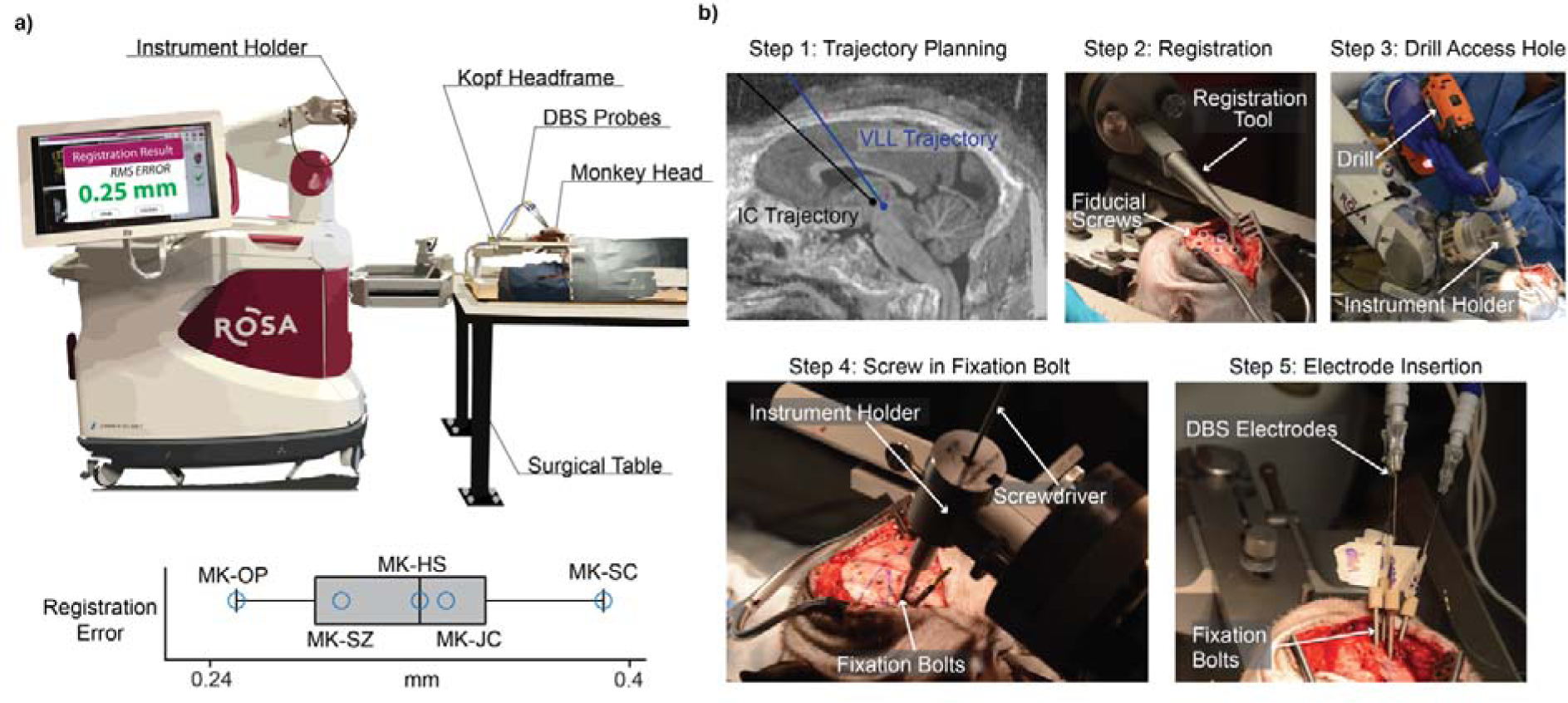
ROSA Setup/Implantation Steps. **(a)** *Top*: Rosa robot surgery setup. This panel was adapted from^68^. *Bottom*: Root mean squared registration error after registration using the fiducial screws for n=5 animals. Co-registration errors were minimal (MK-SC: 0.39 mm, MK-SZ: 0.32 mm, MK-OP: 0.25 mm, MK-HS = 0.29 mm, MK-JC = 0.33 mm), consistent with our previous report^68^, and similar to those observed in human surgeries (0.27 ± 0.07 mm)^80^, thus ensuring highly precise implantation^81^. **(b)** Step 1: we plan the trajectories of each DBS probe in the ROSA One Brain planning software. Step 2: using the Rosa registration tool, we register the position of the brain with fiducial screws in the skull. Step 3: an access hole is drilled into the skull along the trajectory of the probe. Step 4: fixation bolts are screwed into the skull along the probe trajectory. Step 5: DBS and IC electrodes are inserted into fixation bolts at target depth.

**Extended Data Fig. 2:**
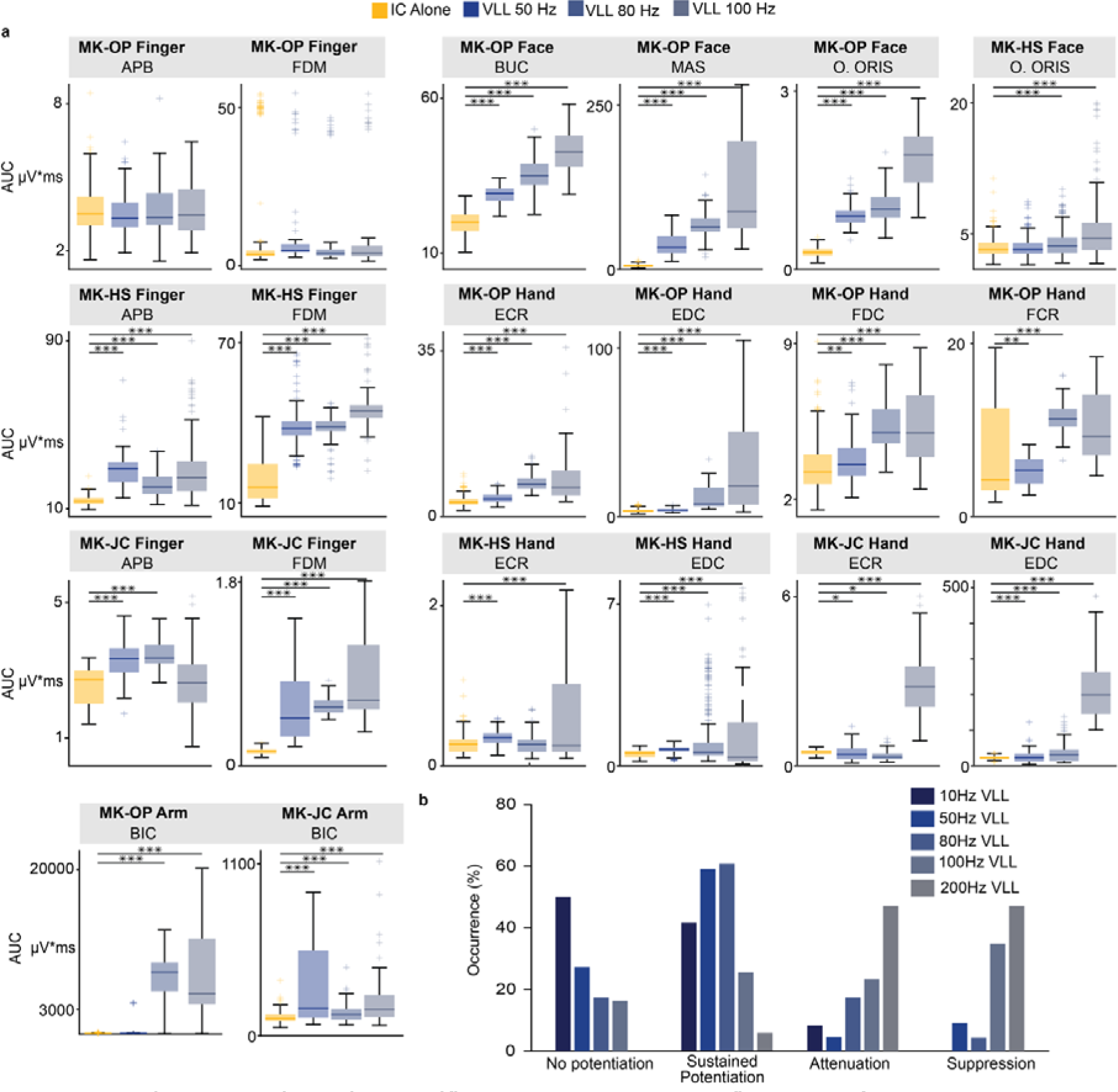
VLL Stimulation amplifies MEP across arm, hand, fingers, and face muscles. **(a)** Boxplots of AUC of MEPs across multiple muscles (APB: Abductor Pollicis Brevis, FDC: Flexor Digitorum Communis, FDM: Flexor Digiti Minimi, EDC: Extensor Digitorum Communis, ECR: Extensor Carpi Radialis, BIC: Biceps, Buc: Buccinator, Mas: Masseter, O. Oris: Orbicularis Oris) in n=3 animals (MK-OP, MK-HS, MK-JC) where EMG recordings were performed. MEPs were recorded during IC stimulation alone (MK-OP: n = 360, MK-HS: n = 237, MK-JC: n = 117) and then paired with VLL stimulation at 50 (MK-OP: n = 68, MK-HS: n = 222, MK-JC: n = 51), 80 (MK-OP: n = 72, MK-HS: n = 233, MK-JC: n = 90), and 100 Hz (MK-OP: n = 63, MK-HS: n = 234, MK-JC: n = 90). Muscles that did not show MEPs responses were not displayed. For all boxplots, the whiskers extend to the maximum spread not considering outliers, central, top, and bottom lines represent median, 25^th^, and 75^th^ percentile, respectively. For all panels, statistical significance was assessed with two-tail bootstrapping with Bonferroni correction: p<0.05 (*), p<0.01 (**), p<0.001(***). **(b)** The occurrence of modulation patterns with respect to stimulation frequency for no potentiation, sustained potentiation, attenuation and suppression.

**Extended Data Fig. 3:**
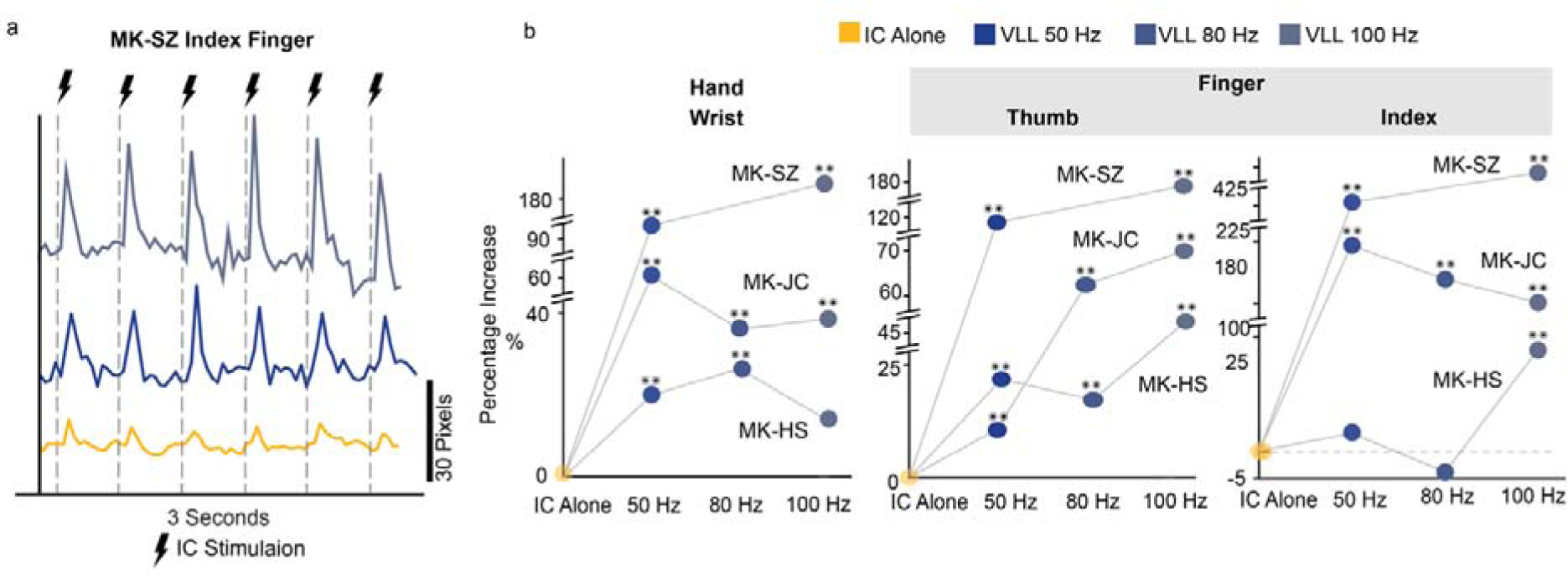
VLL stimulation potentiates movements of the arm and hand. **(a)** Example kinematic trace from MK-SZ with IC alone and paired with VLL stimulation at 50 and 100 Hz. **(b)** Scatter plots for the hand (wrist marker) and fingers (thumb and index marker), representing the percentage of increase of the peak to peak amplitude, in n=3 animals (MK-SZ, MK-HS, MK-JC). Kinematics were recorded during IC stimulation alone and then with paired VLL stimulation at 50, 80, and 100 Hz. For all panels, statistical significance was assessed with two-tail bootstrapping with Bonferroni correction: p<0.05 (*), p<0.01 (**), p<0.001(***).

**Extended Data Fig. 4:**
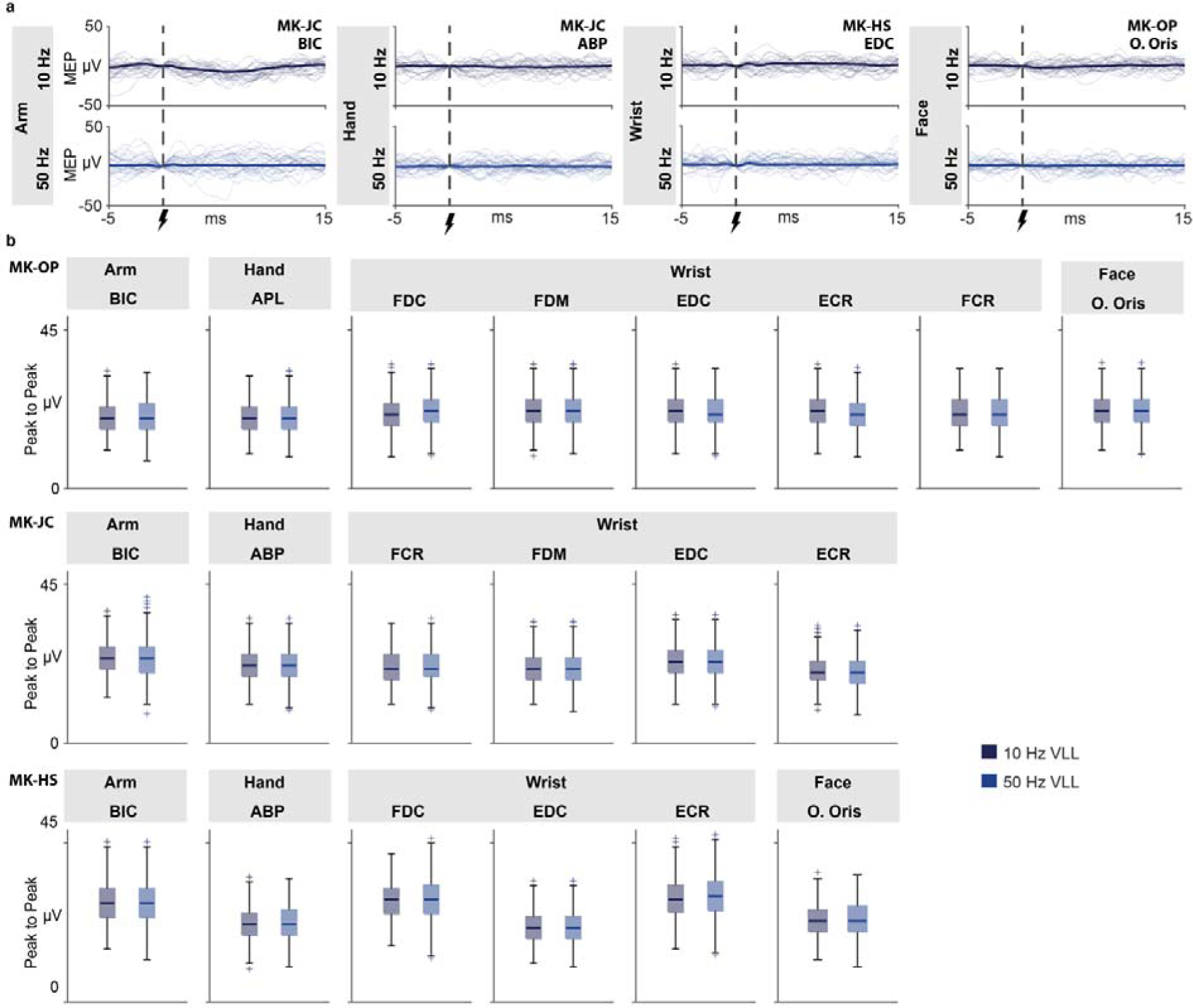
MEP responses from VLL stimulation. **(a)** Example MEPs (30 traces for each plot) of one arm, hand, wrist, and face muscle elicited by VLL stimulation at either 10 Hz (*top row*) or 50 Hz (*bottom row*). **(b)** Boxplots of the AUC MEP responses to VLL stimulation at 10 and 50 Hz for arm (BIC: Bicep), hand (ABP: Abductor Pollicis Brevis and APL: Aductor pollicis longus), wrist (ECR: Extensor Carpi Radialis, EDC: Extensor Digitorum Communis, FCR: Flexor Carpi Radialis, FDC: Flexor digitorum superficialis, and FDM: Flexor digiti minimi brevis), and face (O. Oris: Orbicularis oris) muscles for three animals (MK-OP, MK-JC, MK-HS). For all boxplots, the whiskers extend to the maximum spread not considering outliers, central, top, and bottom lines represent median, 25^th^, and 75^th^ percentile, respectively.

**Extended Data Fig. 5:**
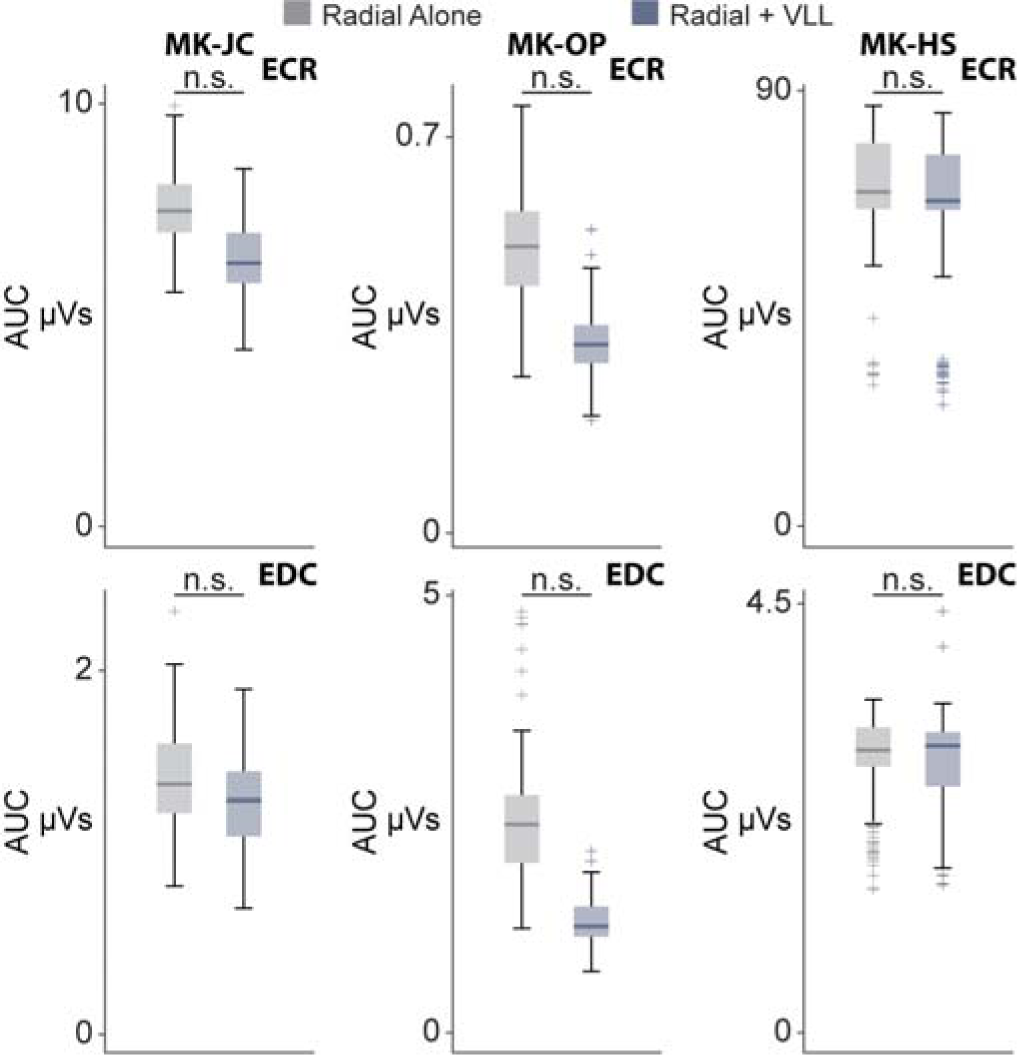
Radial nerve MEPs. Boxplots of the AUC amplitudes of the EMG reflexes for ECR (*top row*) and EDC (*bottom row*) elicited by radial nerve stimulation alone and radial nerve stimulation paired with continuous VLL stimulation at 50 Hz for 3 animals (MK-JC, MK-OP, MK-HS). For all boxplots, the whiskers extend to the maximum spread not considering outliers, central, top, and bottom lines represent median, 25^th^, and 75^th^ percentile, respectively. For all panels, statistical significance was assessed with one-tail bootstrapping with Bonferroni correction, however, in all cases the results were not significant.

**Extended Data Fig. 6:**
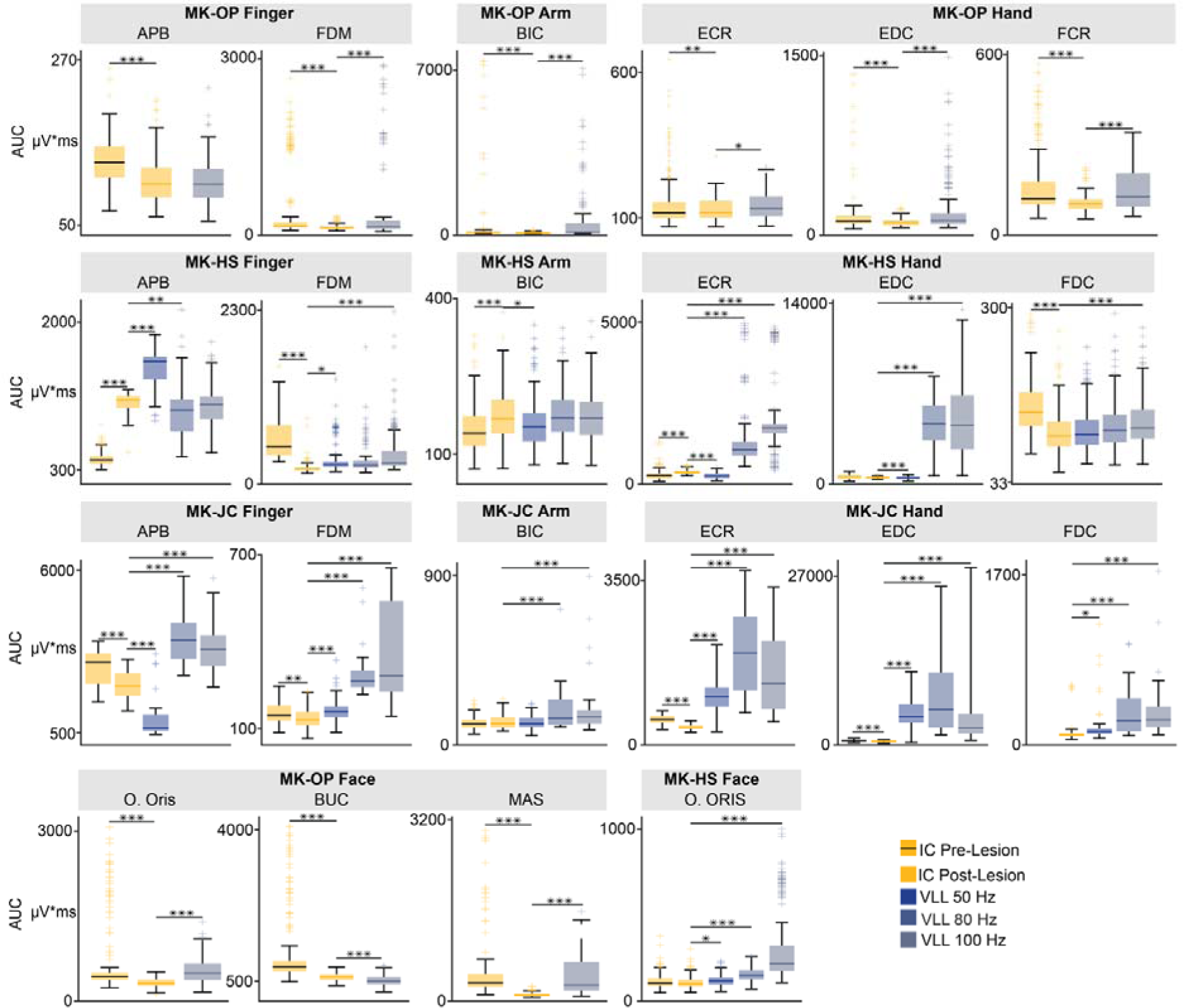
Potentiation of MEP persists after CST lesions across arm, hand, fingers, and face muscles. Boxplots of the AUC of MEPs across multiple muscles (APB: Abductor Pollicis Brevis, FDC: Flexor Digitorum Communis, FDM: Flexor Digiti Minimi, EDC: Extensor Digitorum Communis, ECR: Extensor Carpi Radialis, BIC: Biceps, Buc: Buccinator, Mas: Masseter, O. Oris: Orbicularis Oris) in n=3 animals (MK-OP, MK-HS, MK-JC) where EMG recordings were performed after lesion of the CST. MEPs were recorded during IC stimulation alone before (MK-OP: n = 237, MK-HS: n = 237, MK-JC n = 117) and after the CST lesion (MK-OP: n = 127, MK-HS: n = 169, MK-JC: n = 60) and then paired with VLL stimulation at 50 (MK-HS: n = 172, MK-JC: n = 60), 80 (MK-HS: n = 174, MK-JC: n = 28), and 100 Hz (MK-OP: n = 168, MK-HS: n = 173, MK-JC: n = 60). Muscles that did not show MEPs responses were not displayed. For all boxplots, the whiskers extend to the maximum spread not considering outliers, central, top, and bottom lines represent median, 25^th^, and 75^th^ percentile, respectively. For all panels, statistical significance was assessed with two-tail bootstrapping with Bonferroni correction: p<0.05 (*), p<0.01 (**), p<0.001(***).

**Extended Data Fig.7:**
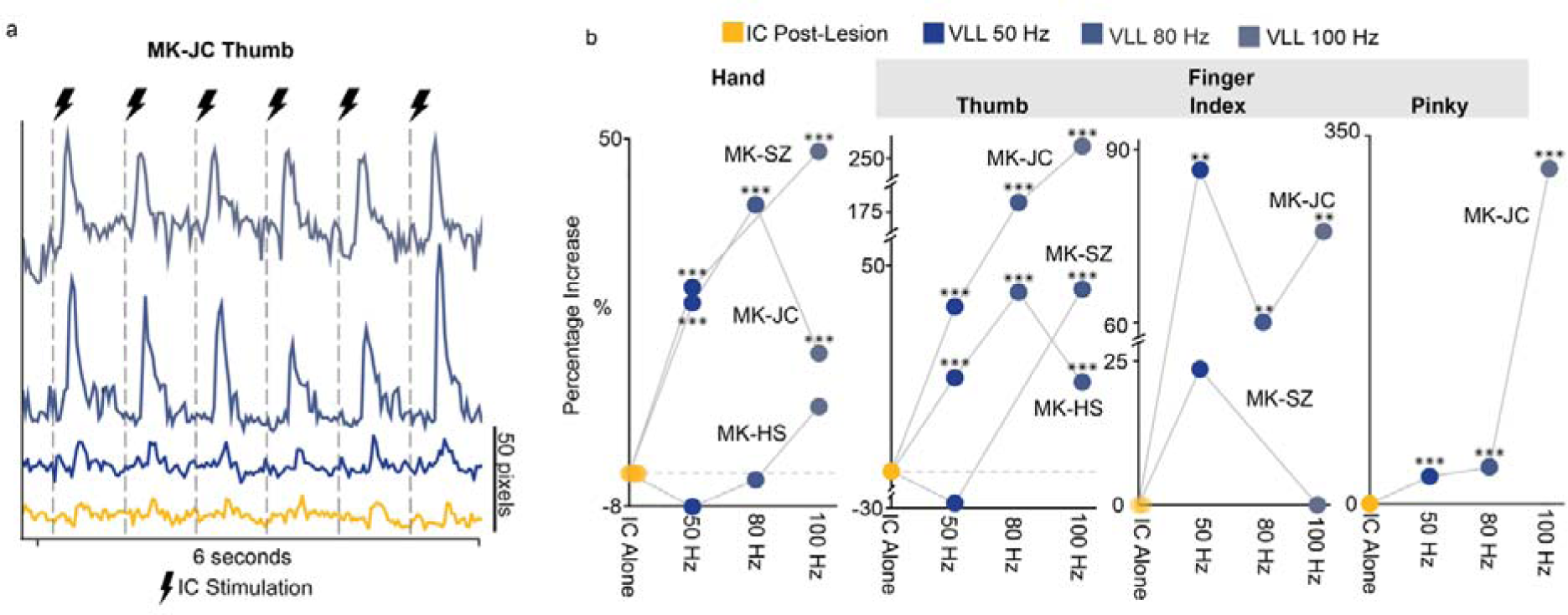
Potentiation of movements persists after CST Lesion. **(a)** Example kinematic trace from MK-JC with IC alone and paired with VLL at 50, 80, and 100 Hz after lesion of the CST. **(b)** Scatter plots for the arm (wrist marker) and hand (thumb, index, and pinky marker), representing the percentage of increase of the peak to peak amplitude, in n=3 animals (MK-SZ, MK-HS, and MK-JC). Traces were created in DeepLabCut with markers placed on the thumb, index finger, pinky finger, and wrist. Kinematics was recorded during IC stimulation alone and IC stimulation paired with VLL stimulation at 50, 80, and 100 Hz.

**Extended Data Fig. 8:**
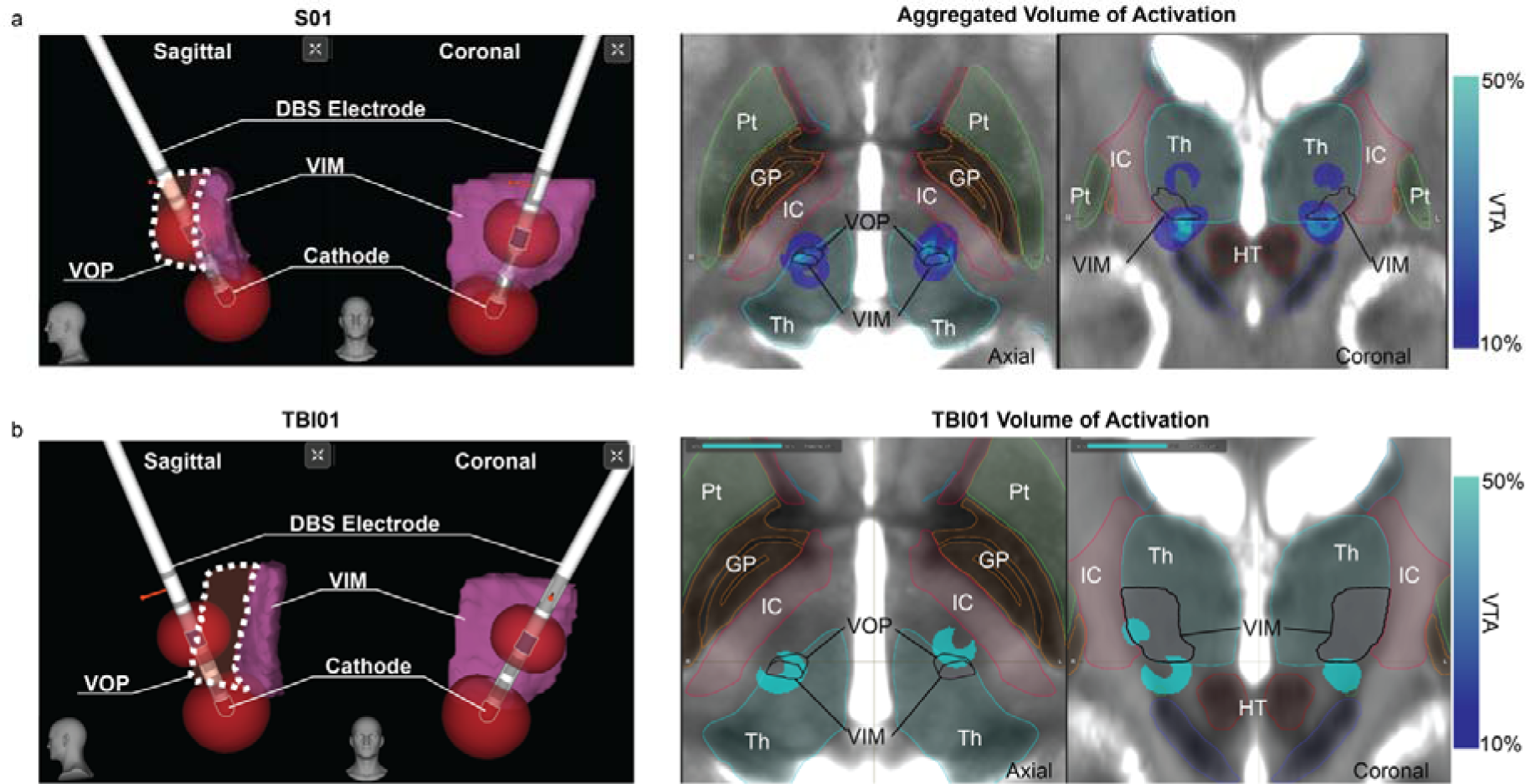
Human DBS Volume of Tissues Activation. **(a)** *Left*: Reconstructions of the VIM/VOP deep brain stimulation (DBS) electrode from S01. Simulated volume of tissue activation (VTA) at the cathode and anode (bipolar stimulation). *Right*: Aggregation of modeled VTAs from n=4 human participants (S01, S02, S03, S04). **(b)** *Left*: Reconstructions of VIM/VOP DBS electrodes and VTA from CST01. *Right*: modeled VTA from VIM/VOP DBS for CST01.

**Extended Data Fig. 9:**
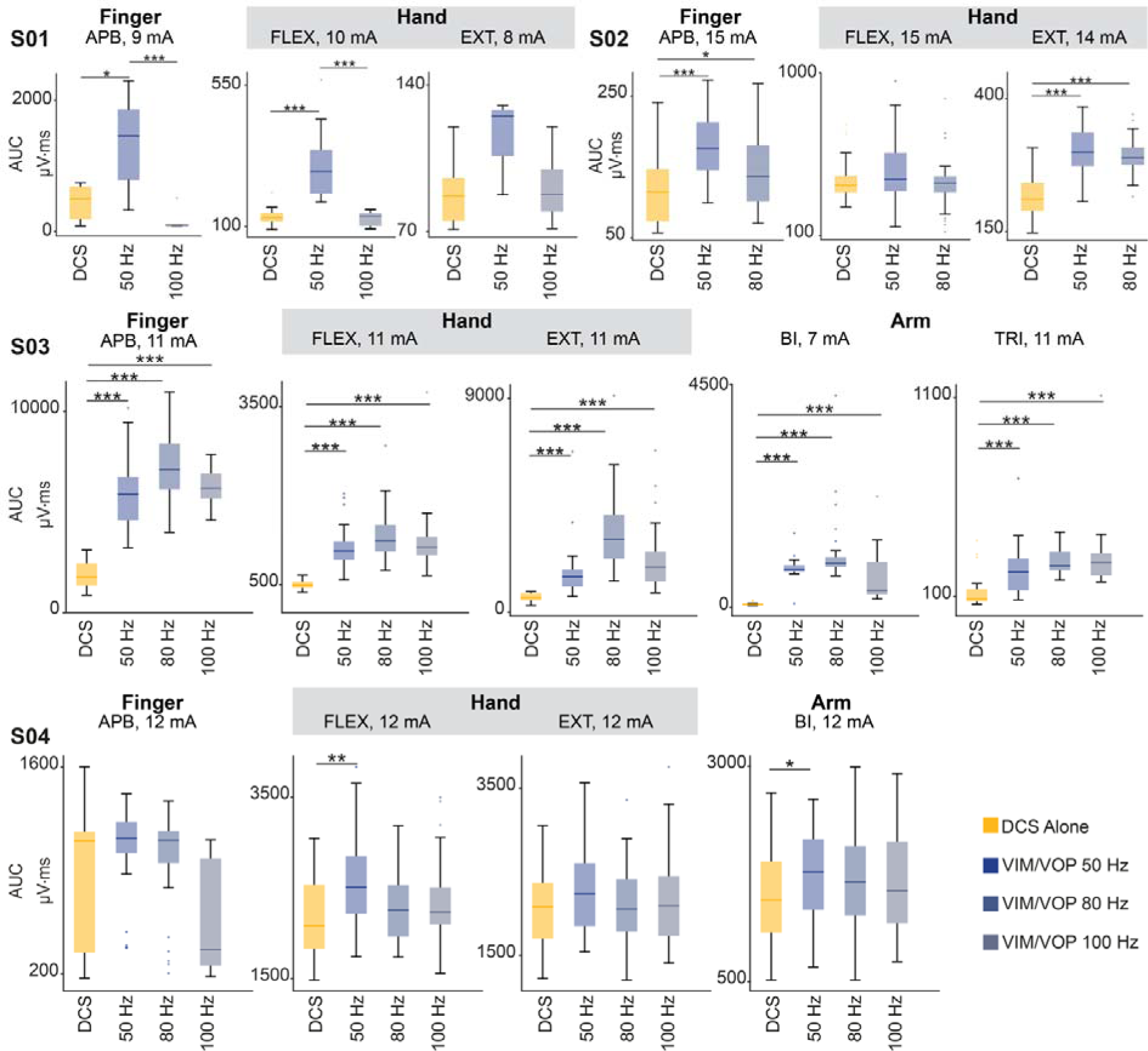
Stimulation of the motor thalamus potentiates MEPs across arm, hand, and fingers muscles in humans. Box plots of MEPs AUC amplitudes of different muscles with DCS alone and DCS paired with VIM/VOP stimulation at 50, and/or 80 Hz and/or 100 Hz. All subjects (S01, S02, S03 and S04) are reported. Amplitudes refer to the current amplitudes for DCS. APB, abductor pollicis brevis, FLEX, flexors; EXT, extensors; BI, biceps; TRI, triceps. For all boxplots, the whiskers extend to the maximum spread not considering outliers, central, top, and bottom lines represent median, 25^th^, and 75^th^ percentile, respectively. For all panels, statistical significance was assessed with two-tail bootstrapping with Bonferroni correction: p<0.05 (*), p<0.01 (**), p<0.001(***).

**Extended Data Fig. 10:**
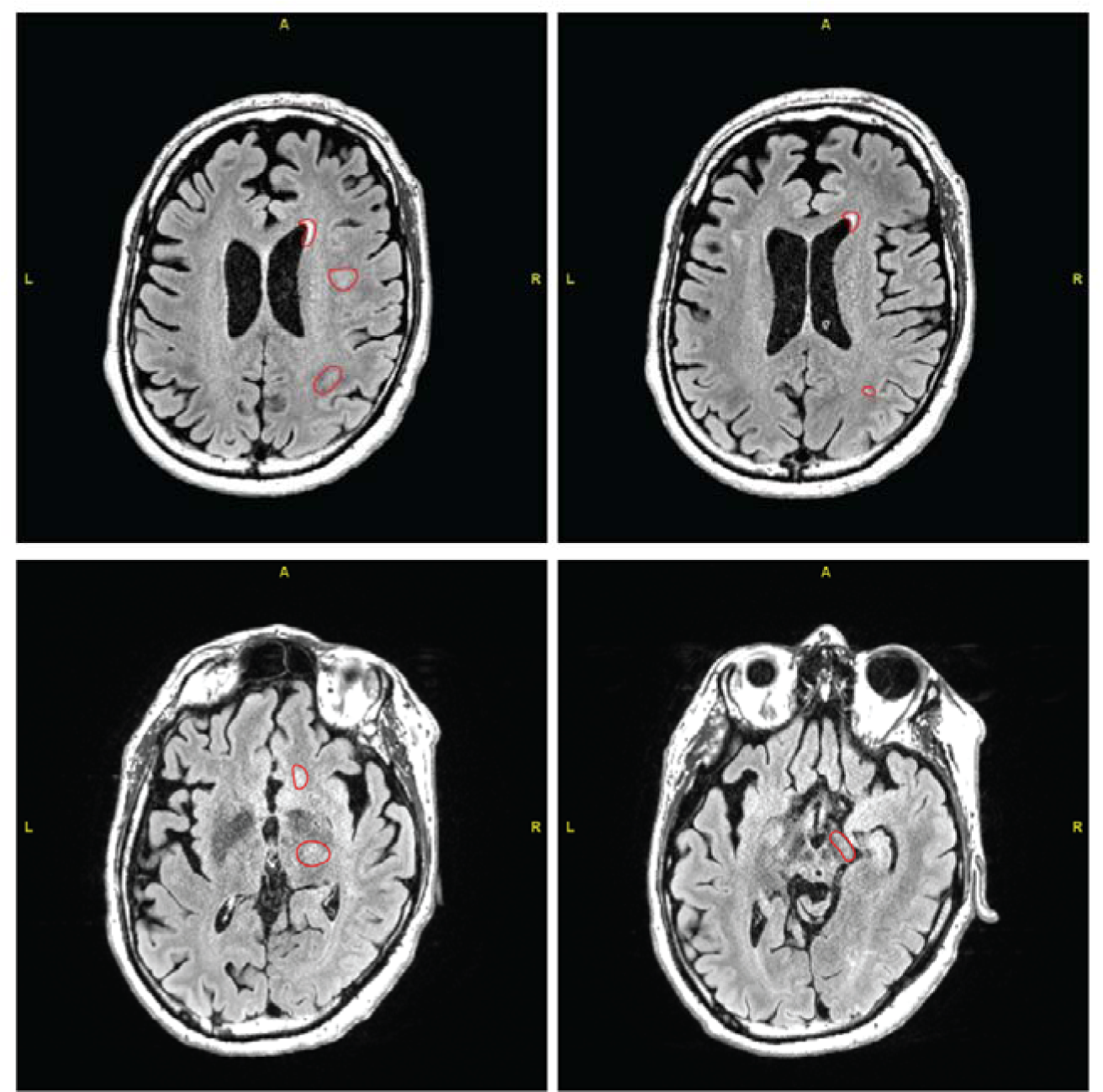
CST01 Lesion Segmentation. Slices of T2-Flair MRI from CST01 with areas of lesion highlighted in red.

**Supplementary Table 1:** Summary of monkey demographic and experiments performed.

| Animal ID | Species | Sex | Age (Yr) | Weight | Experiments |
| --- | --- | --- | --- | --- | --- |
| MK-SC | Macaca Fascicularis | M | 4 | 5.4 kg | Cortical Evoked Potentials/Spike Counts |
| MK-SZ | Macaca Fascicularis | M | 4 | 5.9 kg | Cortical Spike Counts<br>Kinematic Recording (pre/post lesion) |
| MK-OP | Macaca Fascicularis | F | 6 | 6 kg | Cortical Evoked Potentials<br>Antidromic Potentials<br>EMG Recordings (pre/post lesion)<br>Kinematic Recording (pre/post lesion)<br>Radial Nerve Control<br>Frequency Analysis |
| MK-HS | Macaca Mulatta | M | 7 | 12 kg | Cortical Evoked Potentials/Spike Counts<br>Antidromic Potentials<br>EMG Recordings (pre/post lesion)<br>Kinematic Recording (pre/post lesion)<br>Radial Nerve Control<br>Frequency Analysis |
| MK-JC | Macaca Fascicularis | M | 6 | 7.5 kg | Cortical Evoked Potentials<br>Antidromic Potentials<br>EMG Recordings (pre/post lesion)<br>Kinematic Recording (pre/post lesion)<br>Radial Nerve Control<br>Frequency Analysis<br>Force Measurements (pre/post lesion) |

**Supplementary Table 2:**
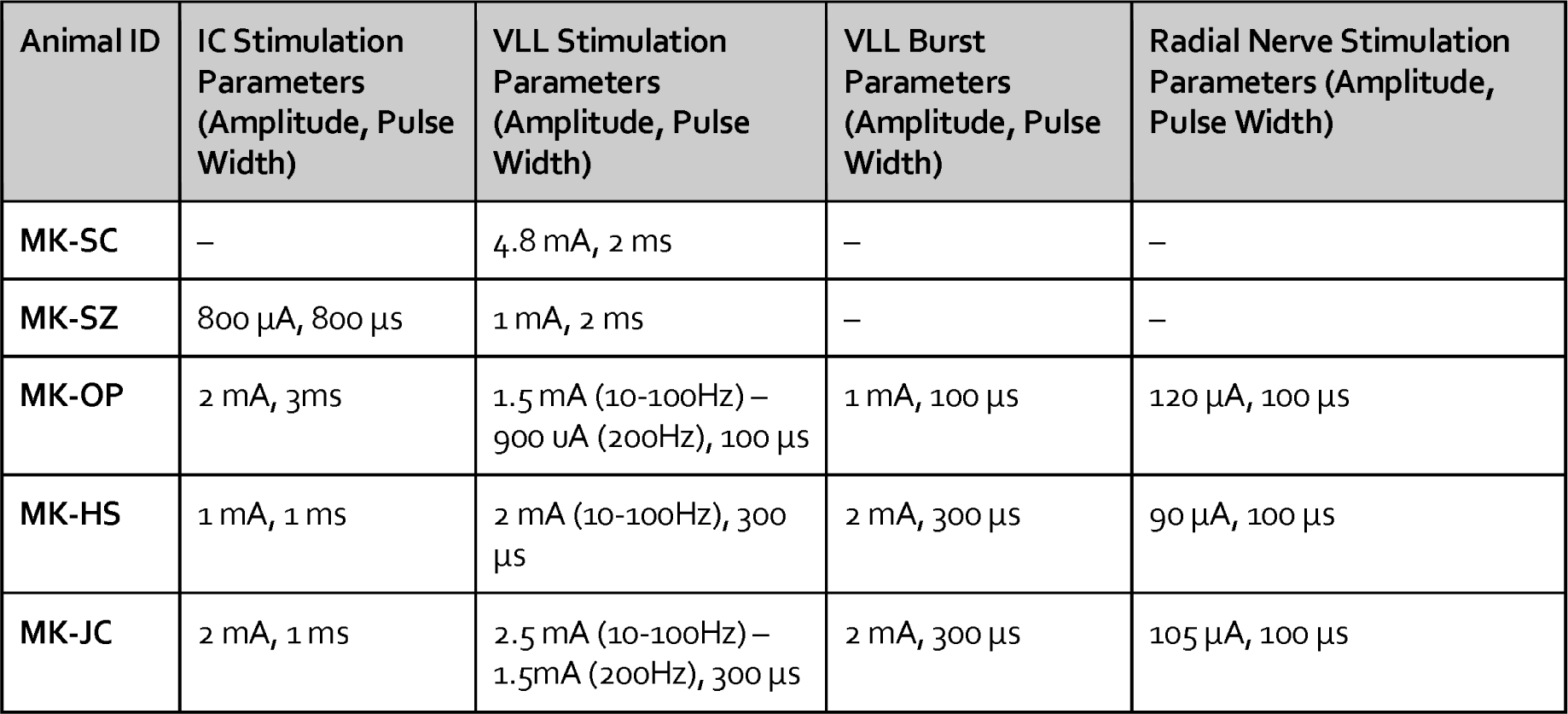
Simulation parameters for IC, VLL, and radial nerve. – indicates that that stimulation was not performed in that particular animal.

**Supplementary Table 3:** Temperature and time used to create a lesion of the CST within the internal capsule.

| Animal ID | Temperature | Time |
| --- | --- | --- |
| **MK-SC** | 90<sup>°</sup> | 60 seconds |
| **MK-SZ** | 75<sup>°</sup> | 60 seconds |
| **MK-OP** | 80<sup>°</sup> | 60 seconds |
| **MK-HS** | 80<sup>°</sup> | 60 seconds |
| **MK-JC** | 80<sup>°</sup> | 30 seconds |

**Supplementary Table 4:** Stimulation parameters for DCS and VIM/VOP in humans. The amplitude of stimulation of the VIM/VOP was always at 3mA and within contact −1 and +8.

| **Subject** | **DCS amplitude** | **VIM/VOP frequency** |
| --- | --- | --- |
| **S01** | 8 mA, 9 mA, 10 mA | 50 Hz, 100 Hz |
| **S02** | 13 mA, 14 mA, 15 mA | 50 Hz, 80 Hz |
| **S03** | 7 mA, 11 mA | 50 Hz, 80 Hz, 100 Hz |
| **S04** | 8 mA, 10 mA, 12 mA | 50 Hz, 80 Hz, 100 Hz |

